# Genome-wide pleiotropy analysis of longitudinal blood pressure and harmonized cognitive performance measures

**DOI:** 10.1101/2025.02.11.25322014

**Authors:** Moonil Kang, Ting Fang Alvin Ang, Sherral A. Devine, Richard Sherva, Shubhabrata Mukherjee, Emily H. Trittschuh, Laura E. Gibbons, Phoebe Scollard, Michael Lee, Seo-Eun Choi, Brandon Klinedinst, Connie Nakano, Logan C. Dumitrescu, Timothy J. Hohman, Michael L. Cuccaro, Andrew J. Saykin, Walter A. Kukull, David A. Bennett, Li-San Wang, Richard P. Mayeux, Jonathan L. Haines, Margaret A. Pericak-Vance, Gerard D. Schellenberg, Paul K. Crane, Rhoda Au, Kathryn L. Lunetta, Jesse Mez, Lindsay A. Farrer

**Affiliations:** Department of Medicine (Biomedical Genetics), Boston University Chobanian & Avedisian School of Medicine, Boston, MA; Department of Anatomy and Neurobiology, Boston University Chobanian & Avedisian School of Medicine, Boston, MA; Framingham Heart Study, Boston University Chobanian & Avedisian School of Medicine, Boston, MA; Slone Epidemiology Center, Boston University Chobanian & Avedisian School of Medicine, Boston, MA; Department of Medicine, University of Washington School of Medicine, Seattle, WA; Geriatric Research, Education, and Clinical Center, Veterans Affairs Puget Sound Health Care System, Seattle, WA; Department of Psychiatry and Behavioral Sciences, University of Washington School of Medicine, Seattle, WA; Vanderbilt Memory & Alzheimer’s Center, Vanderbilt University Medical Center, Nashville, TN; Vanderbilt Genetics Institute, Vanderbilt University Medical Center, Nashville, TN; John P. Hussman Institute for Human Genomics, Miller School of Medicine, Miami, FL; Indiana Alzheimer’s Disease Research Center, Indiana University School of Medicine, Indianapolis, IN; Department of Radiology and Imaging Services, Indiana University School of Medicine, Indianapolis, IN; Department of Medical and Molecular Genetics, Indiana University School of Medicine, Indianapolis, IN; Department of Epidemiology, University of Washington, Seattle, WA; Rush Alzheimer’s Disease Center, Rush University Medical Center, Chicago, IL; Department of Pathology and Laboratory Medicine, University of Pennsylvania Perelman School of Medicine, Philadelphia, PA; Department of Neurology, Columbia University School of Medicine, New York, NY; Cleveland Institute for Computational Biology, Department of Population and Quantitative Health Sciences, Case Western Reserve University, Cleveland, OH; Boston University Alzheimer’s Disease Research Center, Boston University Chobanian & Avedisian School of Medicine, Boston, MA; Department of Epidemiology, Boston University School of Public Health, Boston, MA; Department of Neurology, Boston University Chobanian & Avedisian School of Medicine, Boston, MA; Department of Biostatistics, Boston University School of Public Health, Boston, MA; Department of Ophthalmology, Boston University Chobanian & Avedisian School of Medicine, Boston, MA

**Keywords:** Blood pressure, Cognitive domain score, Alzheimer’s disease, Longitudinal study, Genome-wide association study, Pleiotropy, Differential gene expression, Pathway analysis

## Abstract

**Background:** Genome-wide association studies (GWAS) have identified over 1,000 blood pressure (BP) loci and over 80 loci for Alzheimer’s disease (AD). Considering BP is an AD risk factor, identifying pleiotropy in BP and cognitive performance measures may indicate mechanistic links between BP and AD.

**Methods:** Genome-wide scans for pleiotropy in BP variables—systolic (SBP), diastolic (DBP), mean arterial (MAP), and pulse pressure (PP)—and co-calibrated scores for cognitive domains (executive function, language, and memory) were performed using generalized linear mixed models and 116,075 longitudinal measures from 25,726 participants of clinic-based and prospective cohorts. GWAS was conducted using PLACO to estimate each SNP’s main effect and interaction with age, and their joint effect on pleiotropy. Effects of genome-wide significant (GWS) pleiotropic SNPs on cognition as direct or mediated through BP were evaluated using Mendelian randomization. Potential contribution of genes in top-ranked pleiotropic loci to cognitive resilience was assessed by comparing their expression in brain tissue from pathologically confirmed AD cases with and without clinical symptoms.

**Results:** Pleiotropy GWAS identified GWS associations with *APOE* and 11 novel loci. In the total sample, pleiotropy was identified for SBP and language with *JPH2* (*P*_Joint_=6.09×10^-9^) and *GATA3* (*P*_G×Age_=1.42×10^-8^), MAP and executive function with *PAX2* (*P*_G×Age_=4.22×10^-8^), MAP and language with *LOC105371656* (*P*_G×Age_=1.75×10^-8^), and DBP and language with *SUFU* (*P*_G_=2.10×10^-8^). In prospective cohorts, pleiotropy was found for SBP and language with *RTN4* (*P*_G×Age_=1.49×10^-8^), DBP and executive function with *ULK2* (*P*_Joint_=2.85×10^-8^), PP and memory with *SORBS2* (*P*_G_=2.33×10^-8^), and DBP and memory with *LOC100128993* (*P*_G×Age_=2.81×10^-8^). In clinic-based cohorts, pleiotropy was observed for PP and language with *ADAMTS3* (*P*_G_=2.37×10^-8^) and SBP and memory with *LINC02946* (*P*_G×Age_=3.47×10^-8^). Five GWS pleiotropic loci influence cognition directly, and genes at six pleiotropic loci were differentially expressed between pathologically confirmed AD cases with and without clinical symptoms.

**Conclusion:** Our results provide insight into the underlying mechanisms of high BP and AD. Ongoing efforts to harmonize BP and cognitive measures across several cohorts will improve the power of discovering, replicating, and generalizing novel associations with pleiotropic loci.

## Introduction

Hypertension is a well-established risk factor for dementia [1], particularly Alzheimer’s disease (AD) and vascular dementia—the two most common types of dementia which usually co-exist and collectively account for 85% of dementia cases [2]. Accumulating evidence from large-scale longitudinal studies consistently suggests that elevated blood pressure (BP) in midlife (45−65 years) is associated with a higher risk of late-life dementia [3, 4] and a steeper cognitive decline [5]. However, while the association of late-life high BP with incident AD and AD-related dementias (ADRD) and cognitive decline is controversial in older adults [6], multiple studies report the benefit of lowering BP in elderly hypertensive patients [7, 8] and the association of abnormally lower diastolic BP (DBP) in late-life with a higher risk of cognitive impairment and AD/ADRD [9, 10]. Data from the Framingham Heart Study (FHS) indicate that patterns of age-related systolic BP (SBP) and DBP changes differ in older adults. Compared to SBP which linearly rises across the life course, DBP increases continuously until 50−60 years but decreases after age 60, corresponding to a late-life increase in pulse pressure (PP) [11].

Although multiple genome-wide association studies (GWAS) have identified more than 1,000 loci for various BP traits [12-14] and 80 loci associated with the risk of AD/ADRD [15-18], only a few studies have investigated their shared genetic architecture [19, 20] that has been implied by neuropathological evidence showing a link between high BP and AD [21-23]. Here, we investigated pleiotropy, occurring when a single gene or variant affects two or more phenotypes [24], for BP traits and measures of cognitive performance on a genome-wide scale in several large clinic-based and prospective cohorts. Identifying pleiotropy for BP and cognitive performance measures may provide more insight into the genetic basis and mechanistic links between high BP and AD/ADRD. In addition, based on recent evidence suggesting that BP and biomarkers of vascular integrity may impact cognitive resilience through tau pathology [25, 26], we compared expression of genes located within top-ranked loci from the GWAS in brain tissue obtained from individuals with pathologically confirmed AD who were cognitively impaired or normal (*i*.*e*., cognitively resilient) prior to death to test the hypothesis that some of these pleiotropic genes may influence propensity to cognitive resilience in addition to or independent of mechanisms leading to AD hallmark amyloid-β (Aβ) and tau pathologies.

## Methods

### Participant ascertainment and assessment

This study included non-Hispanic white participants aged 60 or older from five longitudinal cohort studies. Three cohorts, including the FHS [27-29], the Adult Changes in Thought (ACT) Study [30], and the Religious Orders Study/Rush Memory and Aging Project (ROSMAP) [31], recruited cognitively normal participants and followed them over time. The National Institute on Aging (NIA)-sponsored Alzheimer’s Disease Research Centers, which collect a uniform set of phenotypic data archived by the National Alzheimer’s Coordinating Center (NACC) [32, 33], and the Alzheimer’s Disease Neuroimaging Initiative (ADNI) [34, 35] are clinic-based cohorts. Individuals in these cohorts included in this study were cognitively normal or met the criteria for mild cognitive impairment (MCI) or AD at the most recent examination. All participants underwent an examination at enrollment and each follow-up visit, including basic anthropometry, BP measurements, medications, and medical history. For those taking antihypertensive medications at the time point when BP was measured, we added 10 and 5 mmHg to the measured SBP and DBP, respectively, as previously recommended [36]. These corrected SBP and DBP were used to calculate PP (SBP minus DBP) and mean arterial pressure (MAP; DBP plus one third of PP) [37]. Body mass index (BMI) was calculated in the unit of kg/m^2^ as one’s weight divided by the square of height at each examination. Further information regarding the ascertainment, evaluation, and diagnostic procedures in each cohort was described elsewhere [30-32, 35, 38, 39].

### Cognitive domain scores

As described previously [40-42], cognitive domain scores for executive function, language, and memory were calculated from tests from unique cognitive batteries administered across the five cohort studies and co-calibrated to be on the same scale across cohorts. Briefly, an expert panel of neuropsychologists and behavioral neurologists classified neuropsychological (NP) test items into one of the cognitive domains. Anchor items, identical NP test items found in multiple batteries, were used to put composite scores on the same scale across studies. Co-calibrated composite scores for each cognitive domain were generated using confirmatory factor analysis in Mplus [43] with loadings for anchor items forced to be equal across studies. Co-calibrated cognitive scores with a standard error (SE) > 0.6 and those obtained solely from the Mini-Mental State Examination (MMSE), which shows a ceiling effect [42], were excluded. We also excluded data collected at ages < 60 that were available only for FHS participants to mitigate the concern that the genetic architecture of BP and/or cognitive performance may differ at younger ages.

### Genotype QC, generating PC, and GRM

Genotype quality control (QC) procedures were applied to the Trans-Omics for Precision Medicine (TOPMed)-imputed genome-wide single nucleotide polymorphism (SNP) data [18, 44], which were aligned to the Genome Research Consortium human build 38 (GRCh38). After excluding variants with poor imputation quality (*r*^2^ < 0.3) or minor allele frequency (MAF) < 0.01, roughly 8.7 million variants remained for each individual cohort. We performed linkage disequilibrium (LD) pruning for genotyped variants (MAF > 0.05 and call rate > 99%) with an LD threshold of 0.1, and principal components (PCs) of the population structure for each sample within each cohort study were derived from the LD-pruned variants using GENESIS [45]. A kinship matrix for family-based samples was estimated for FHS participants using self-reported pedigree information and the R package kinship2 [46]. An empirical genetic relationship matrix (GRM) was derived for other individuals using established procedures [47-49].

### GWAS for BP and cognitive domain measures

We performed GWAS for SBP, DBP, MAP, and PP in each dataset separately using the GMMAT [50] and MAGEE [51] software. The association of each SNP with BP traits over time was evaluated in the following generalized linear mixed model:

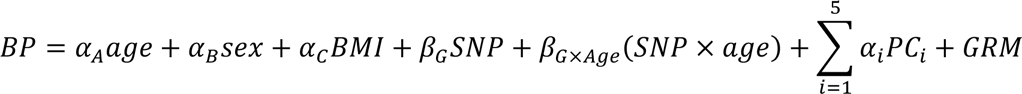

In the model, *α_A_*, *α_B_*, and *α_C_* represent the effects of age, sex, and BMI, respectively, and *α_i_* denotes the effect of the first five PCs. *β_G_* and *β_G×Age_* indicate the SNP and SNP×age interaction effects, respectively. GRM was also included in the model as a random effect. We utilized the GWAS results for executive function, language, and memory that were generated previously in each dataset by applying similar models including the SNP and SNP×age interaction terms, and covariates for age, sex, educational level (less than high school, high school, some college, or college graduate), GRM, and the first five PCs [52]. The SNP×age interaction term accounts for the inclusion of data collected from individuals at multiple time points and allows for the possibility of associations that are age-dependent. In models for all traits, age was centered by subtracting the median age for all observations for all individuals in the dataset from the age at each exam to alleviate the multicollinearity and improve the interpretability of the SNP×age interaction model coefficients [53]. A random age slope and intercept were incorporated in the model to account for repeated measures. Results from each dataset were combined by meta-analysis using the inverse variance-weighted method in METAL [54]. We also calculated a *P*-value for the joint test of the null hypothesis that the SNP and SNP×age interaction effects are both zero by combining *Z*-scores for the SNP’s main effect and SNP×age interaction. To reduce systematic inflation caused by jointly testing the SNP’s main and SNP×age interaction effects [55, 56], we applied a joint meta-analysis approach that takes account of the covariance between their coefficients [57]. Results were also combined in clinic-based cohorts (NACC and ADNI) and prospective cohorts (FHS, ACT, and ROSMAP) separately to allow for unique associations due to the disparity in age and AD ascertainment between these groups (**Figure S1**) [52]. *P*-values were further corrected by applying the genomic inflation factor (*λ*) estimated for each GWAS and were considered genome-wide significant (GWS) if the corrected *P*-value was less than 5×10^-8^.

### Genetic correlations and genome-wide pleiotropy analyses

Genetic correlations among the BP and cognitive domain measures were estimated by cross-trait LD score regression [58, 59] using meta-analyzed GWAS summary statistics for each trait and LD scores derived previously from European ancestry samples in the 1000 Genomes Project Phase 3 data. We performed genome-wide pleiotropy analyses for paired outcomes of BP and cognitive domain traits by combining summarized GWAS results for the individual traits in the total sample and separately in the clinic-based and prospective cohorts (**Figure S1**). Using the R package PLACO [60], we examined the composite null hypothesis that a maximum of one phenotype is associated with a given variant such that rejecting this hypothesis implies that the variant affects both phenotypes and is thus pleiotropic [24]. The PLACO test statistic is the product of the *Z*-scores for a given variant estimated from GWAS for each individual phenotype and follows a mixture distribution that allows for the variant to be associated with at most one phenotype [60]. Potential type I error rate inflation due to *Z*-scores derived from GWAS for BP and cognitive measures with overlapping samples between two phenotypes [61] was corrected based on the Pearson correlation among *Z*-scores for variants with no associations with any trait (*P* > 1×10^-4^), as recommended [60]. Variants with extreme effects (squared *Z*-score > 80) for any trait, which could indicate spurious signals of pleiotropy [58, 62], were excluded.

### Differentiation of directional pleiotropy from mediated pleiotropy

For top-ranked GWS pleiotropic loci, we conducted Mendelian randomization (MR) using the Egger method [63] to differentiate directional pleiotropy (*i*.*e*., SNP’s direct effect on cognition or the effect via confounders other than BP) from mediated pleiotropy (*i*.*e*., SNP’s effect on cognition through BP). The regression model tested for each locus included the most significant GWS pleiotropic variant and correlated proximal SNPs in LD as instrumental variables. The MR-Egger model was evaluated using GWAS summary statistics for the instrumental variable SNPs and the R package MendelianRandomization [64].

### Differential gene expression analyses

We compared the expression of genes located at top-ranked pleiotropic loci among cognitively normal controls (*i*.*e*., neither clinically nor pathologically diagnosed with AD), pathologically confirmed asymptomatic AD cases (*i*.*e*., cognitively normal prior to death and thus considered to be resilient), and pathologically confirmed symptomatic AD cases (*i*.*e*., cognitively impaired or demented prior to death). Genes containing or adjacent (closer than 100 kb) to variants showing GWS evidence of pleiotropy in the SNP main effect, SNP interaction with age, or their joint effects in the total, clinic-based, or prospective cohort samples were targeted. We obtained bulk RNA-sequencing data measured in the dorsolateral prefrontal cortex (DLPFC) from ROSMAP (195 controls, 172 asymptomatic AD, and 200 symptomatic AD cases) [65], Boston University Alzheimer’s Disease Research Center (BUADRC) (35 controls, 20 asymptomatic AD, and 30 symptomatic AD cases) [66], and FHS (73 controls, 12 asymptomatic AD, and 42 symptomatic AD cases) [67] participants. Results were adjusted for a false discovery rate (FDR) by applying the Benjamini-Hochberg method [68] and considered significant if the FDR was less than a threshold of 0.05.

### Pathway enrichment analyses

We conducted pathway enrichment analyses, each of which was seeded with genes containing variants showing evidence of pleiotropy (*P* < 1×10^-4^) in the total sample, clinic-based cohorts, or prospective cohorts using the Ingenuity Pathway Analysis software (QIAGEN Inc.) [69]. The Benjamini-Hochberg method [68] was applied to adjust each canonical pathway’s enrichment *P*-value in each sample stratum for each pair of traits. Pathways were considered significant if the FDR was less than a threshold of 0.05.

## Results

### Basic characteristics

Data available for analyses were obtained from 116,075 longitudinal examinations of 25,726 participants with a mean age of 76.5 years, more than half of whom were female (55.4%) and college graduates (59.5%) (**Tables 1 and S1**). Even though FHS participants were much younger (*P* < 0.001) than those in the other cohorts (70.5 *vs*. 76.5−81.9 years), the mean SBP and PP were significantly higher (*P* < 0.001) in FHS participants compared to individuals in NACC, ADNI, and ROSMAP (139.9 *vs*. 136.2−138.9 mmHg for SBP; 63.5 *vs*. 61.6−62.2 mmHg for PP), which may be because FHS included untreated hypertensive participants who were recruited longer ago. Participants in the clinic-based cohorts (NACC and ADNI) were slightly younger at the last visit than those in the prospective cohorts (FHS, ACT, and ROSMAP). Compared to prospective cohorts, individuals in the clinic-based cohorts had a significantly higher proportion of MCI or AD cases (*P* < 0.001), lower scores for executive function and memory, and higher scores for language (*P* < 0.001) at the last visit [52].

**Table 1.** Characteristics of the study sample.

|  | Total sample | Clinic-based cohorts |  |  | Prospective cohorts |  |  |  |
| --- | --- | --- | --- | --- | --- | --- | --- | --- |
|  |  | NACC | ADNI | Combined | FHS | ACT | ROSMAP | Combined |
| Observations, n | 116,075 | 59,862 | 6,824 | 66,686 | 19,820 | 13,692 | 15,877 | 49,389 |
| Participants, n | 25,726 | 14,360 | 1,335 | 15,695 | 4,976 | 3,009 | 2,046 | 10,031 |
| Age, years (mean±SD) | 76.5±8.5 | 76.5±8.2 | 76.8±7.0 | 76.5±8.1 | 70.5±7.7 | 78.8±7.0 | 81.9±7.2 | 76.5±8.9 |
| Sex, female, n (%) | 14,256 (55.4) | 7,854 (54.7) | 583 (43.7) | 8,437 (53.8) | 2,703 (54.3) | 1,686 (56.0) | 1,430 (69.9) | 5,819 (58.0) |
| Educational level, n (%)<sup>†</sup> |  |  |  |  |  |  |  |  |
| Under high school degree | 974 (4.2) | 302 (2.3) | 41 (3.0) | 343 (2.4) | 305 (8.5) | 240 (7.9) | 86 (4.2) | 631 (7.2) |
| High school degree | 3,987 (17.3) | 1,958 (15.1) | 160 (11.7) | 2,118 (14.8) | 927 (25.7) | 620 (20.4) | 322 (15.5) | 1,869 (21.5) |
| Some college | 4,387 (19.0) | 2,212 (17.0) | 259 (18.9) | 2,471 (17.2) | 905 (25.1) | 688 (22.7) | 323 (15.6) | 1,916 (22.0) |
| Over college graduate | 13,717 (59.5) | 8,513 (65.6) | 907 (66.3) | 9,420 (65.6) | 1,470 (40.8) | 1,487 (49.0) | 1,340 (64.7) | 4,297 (49.3) |
| BMI, kg/m<sup>2</sup> (mean±SD) | 26.7±4.7 | 26.5±4.7 | 26.5±4.5 | 26.5±4.6 | 27.3±4.8 | 26.8±4.6 | 27.0±5.1 | 27.1±4.9 |
| BP, mmHg (mean±SD) |  |  |  |  |  |  |  |  |
| SBP | 139.2±20.0 | 138.9±19.5 | 136.2±18.0 | 138.7±19.4 | 139.9±20.8 | 142.7±21.8 | 137.6±19.4 | 139.9±20.7 |
| DBP | 76.2±10.6 | 76.7±10.4 | 74.6±10.0 | 76.5±10.4 | 76.4±10.5 | 75.0±10.8 | 75.5±10.9 | 75.7±10.8 |
| MAP | 97.2±12.0 | 97.5±11.8 | 95.2±11.0 | 97.2±11.7 | 97.5±12.2 | 97.6±12.9 | 96.2±12.2 | 97.1±12.4 |
| PP | 63.0±16.8 | 62.2±16.5 | 61.6±15.4 | 62.2±16.4 | 63.5±17.8 | 67.7±17.8 | 62.0±15.8 | 64.2±17.3 |
| Cognitive score (mean±SD)<sup>†</sup> |  |  |  |  |  |  |  |  |
| Executive function | 0.197±0.752 | 0.163±0.827 | 0.318±0.746 | 0.182±0.819 | -0.007±0.688 | 0.189±0.499 | 0.343±0.698 | 0.217±0.650 |
| Language | 0.353±0.808 | 0.472±0.869 | 0.392±0.700 | 0.463±0.851 | 0.229±0.662 | 0.166±0.422 | 0.154±0.805 | 0.173±0.695 |
| Memory | 0.337±0.890 | 0.372±1.007 | 0.146±0.873 | 0.346±0.995 | 0.286±0.636 | 0.512±0.549 | 0.194±0.836 | 0.324±0.721 |
| Cognitive status, n (%)<sup>†</sup> |  |  |  |  |  |  |  |  |
| MCI | 2,247 (9.7) | 820 (6.3) | 444 (32.5) | 1,264 (8.8) | 287 (8.0) | NA | 696 (33.6) | 983 (11.3) |
| AD | 9,272 (40.2) | 7,090 (54.6) | 486 (35.6) | 7,576 (52.8) | 445 (12.3) | 485 (16.0) | 766 (37.0) | 1,696 (19.5) |
| Dementia (other than AD) | 627 (2.7) | 184 (1.4) | NA | 184 (1.3) | 135 (3.7) | 283 (9.3) | 25 (1.2) | 443 (5.1) |

<sup>†</sup>Characteristics were based on individuals included in the GWAS for cognitive domain scores.

### Phenotypic and genetic correlations

As shown in **Figure S2**, phenotypic and genetic correlations (*r*) were moderate to high for all pairs of the four BP measures (phenotypic *r*=0.51−0.88, genetic *r*=0.43−0.91), except for the DBP and PP (phenotypic *r*=0.05, genetic *r*=0.03), and for all pairs of the three cognitive domains (phenotypic *r*=0.72−0.78, genetic *r*=0.55−0.73). Predictably, SBP, MAP, and PP were negatively correlated with cognitive domain scores consistently, even though they had weak phenotypic and genetic correlations ranging from -0.16 to -0.04 and -0.07 to -0.01, respectively.

### Genetic associations with individual BP and cognitive domain scores

We identified six GWS BP loci, including *SLC7A1*, *ULK4*, *HOTTIP*, *IGFBP3*, *PIK3CG*, and *MCTP2* (**Table S2a**), with little evidence of genomic inflation (*λ*=1.017−1.076) in the joint testing of the SNP and SNP×age interaction effects across all sample strata (**Figure S3**). GWS associations of top-ranked SNPs in each BP locus were primarily due to the effect of the SNP rather than the SNP×age interaction and were supported by several adjacent variants in high LD (**Figure S4 and Table S3**). Previously, we identified GWS associations of individual cognitive domain scores with *ULK2*, *CDK14*, *LINC02712*, *PURG*, and several established AD loci (*BIN1*, *CR1*, *MS4A6A*, and *GRN*) (**Table S2b**) in addition to GWS associations of many SNPs in the *APOE* region with all cognitive domains [52].

### Pleiotropy for BP and cognitive domain scores

We identified GWS pleiotropic associations with SNPs in *JPH2* and *ULK2* emerging from the total sample and prospective cohorts, respectively (**Tables 2 and S4**), with no evidence of genomic inflation (*λ*=0.868−0.968) in the joint testing of the main SNP and SNP×age interaction effects on pleiotropy between BP and cognitive domain scores across all strata of the sample (**Figure S5**). The *JPH2* association observed between SBP paired with language and rs6031436 (*P*_Joint_=6.09×10^-9^) in the total sample (**Table 2**) was supported by multiple GWS SNPs in the same locus (**Figure 1a and Table S5**). This *JPH2* SNP also showed a GWS pleiotropy for PP paired with language (*P*_Joint_=3.25×10^-8^) in the total sample (**Table 2**), which was supported by multiple adjacent variants in high LD (**Figure 1b and Table S5**). In the prospective cohorts, a GWS pleiotropy was observed with *ULK2* SNP rs157398 (*P*_Joint_=2.85×10^-8^) for the pair of DBP and executive function (**Table 2**), which was supported by many neighboring GWS or suggestive variants (**Figure 2a and Table S5**). In addition, many SNPs in the *APOE* region showed GWS pleiotropy for all the pairs of BP and cognitive performance measures across all strata of analyses, particularly in the clinic-based cohorts (**Table S5**).

**Figure 1.**
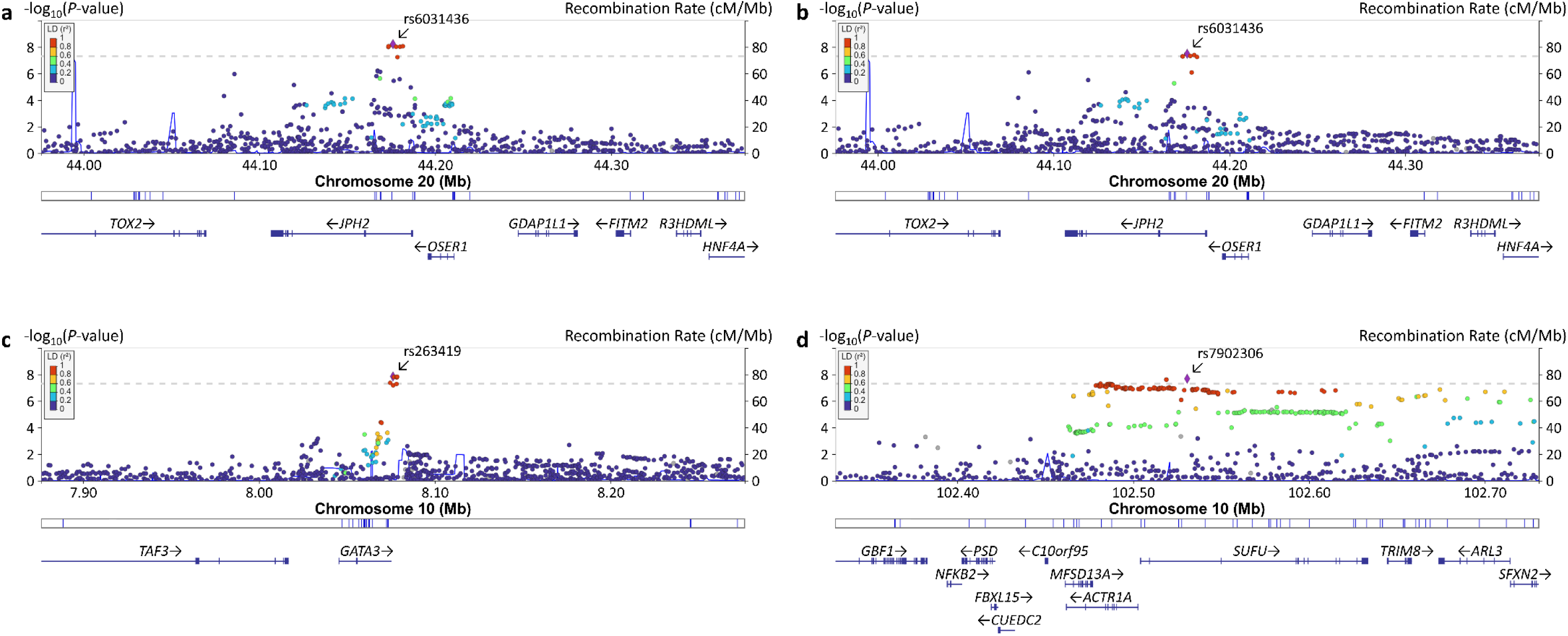
LocusZoom plots for top-ranked pleiotropic loci emerging from the total sample. (a) rs6031436 and *JPH2* variants showing pleiotropy between SBP and language in the total sample, (b) rs6031436 and *JPH2* variants showing pleiotropy between PP and language in the total sample, (c) rs263419 and *GATA3* variants showing pleiotropy between SBP and language in the total sample, and (d) rs7902306 and *SUFU* variants showing pleiotropy between DBP and language in the total sample. A purple diamond indicates the top-ranked SNP at each locus. SNPs are color-coded according to their LD (*r*^2^) with the top-ranked SNP in the region. Horizontal dotted lines represent the GWS threshold of *P*=5×10^-8^. Vertical blue lines indicate locations of the high recombination rate among SNPs at the chromosomal position. Approximate location, transcription direction, and coding portions (exons represented by vertical bars) of genes are shown below the diagram. Mb=megabase, cM=centimorgan.

**Figure 2.**
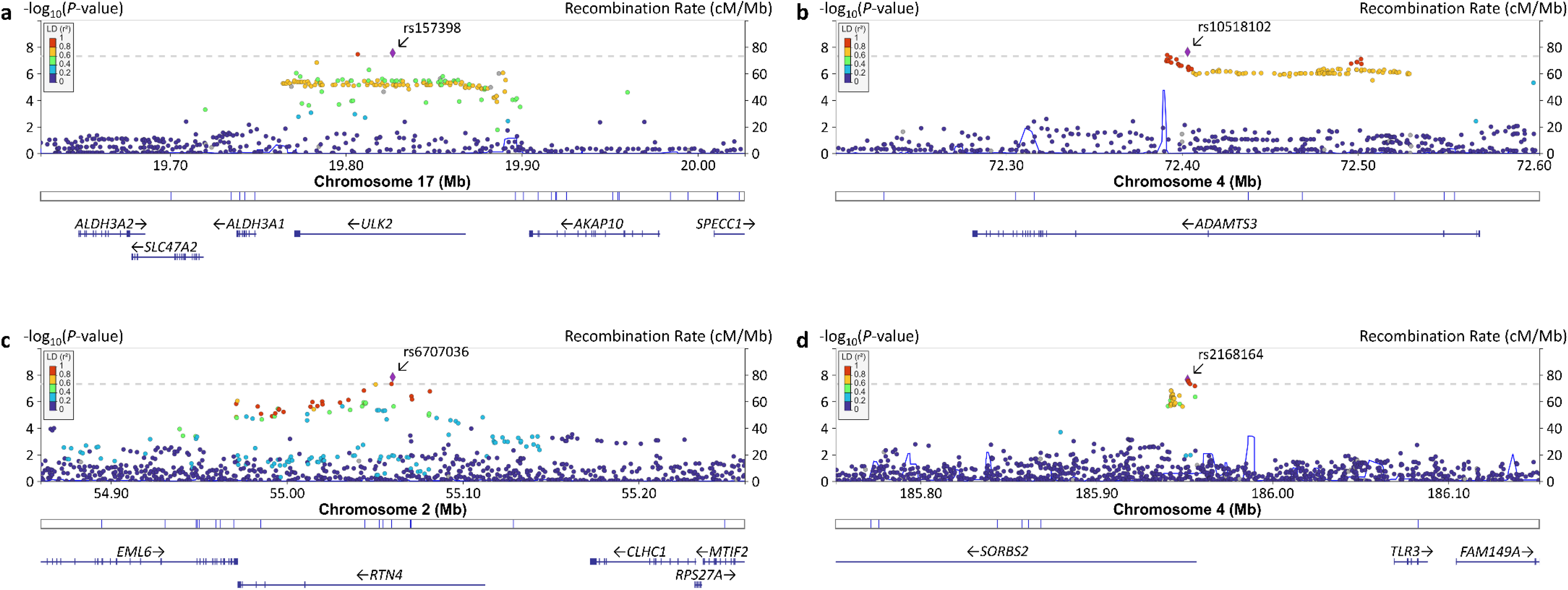
LocusZoom plots for top-ranked pleiotropic loci emerging from the clinic-based or prospective cohort samples. (a) rs157398 and *ULK2* variants showing pleiotropy between DBP and executive function in the prospective cohorts, (b) rs10518102 and *ADAMTS3* variants showing pleiotropy between PP and language in the clinic-based cohorts, (c) rs6707036 and *RTN4* variants showing pleiotropy between SBP and language in the prospective cohorts, and (d) rs2168164 and *SORBS2* variants showing pleiotropy between PP and memory in the prospective cohorts. A purple diamond indicates the top-ranked SNP at each locus. SNPs are color-coded according to their LD (*r*^2^) with the top-ranked SNP in the region. Horizontal dotted lines represent the GWS threshold of *P*=5×10^-8^. Vertical blue lines indicate locations of the high recombination rate among SNPs at the chromosomal position. Approximate location, transcription direction, and coding portions (exons represented by vertical bars) of genes are shown below the diagram. Mb=megabase, cM=centimorgan.

**Table 2.**
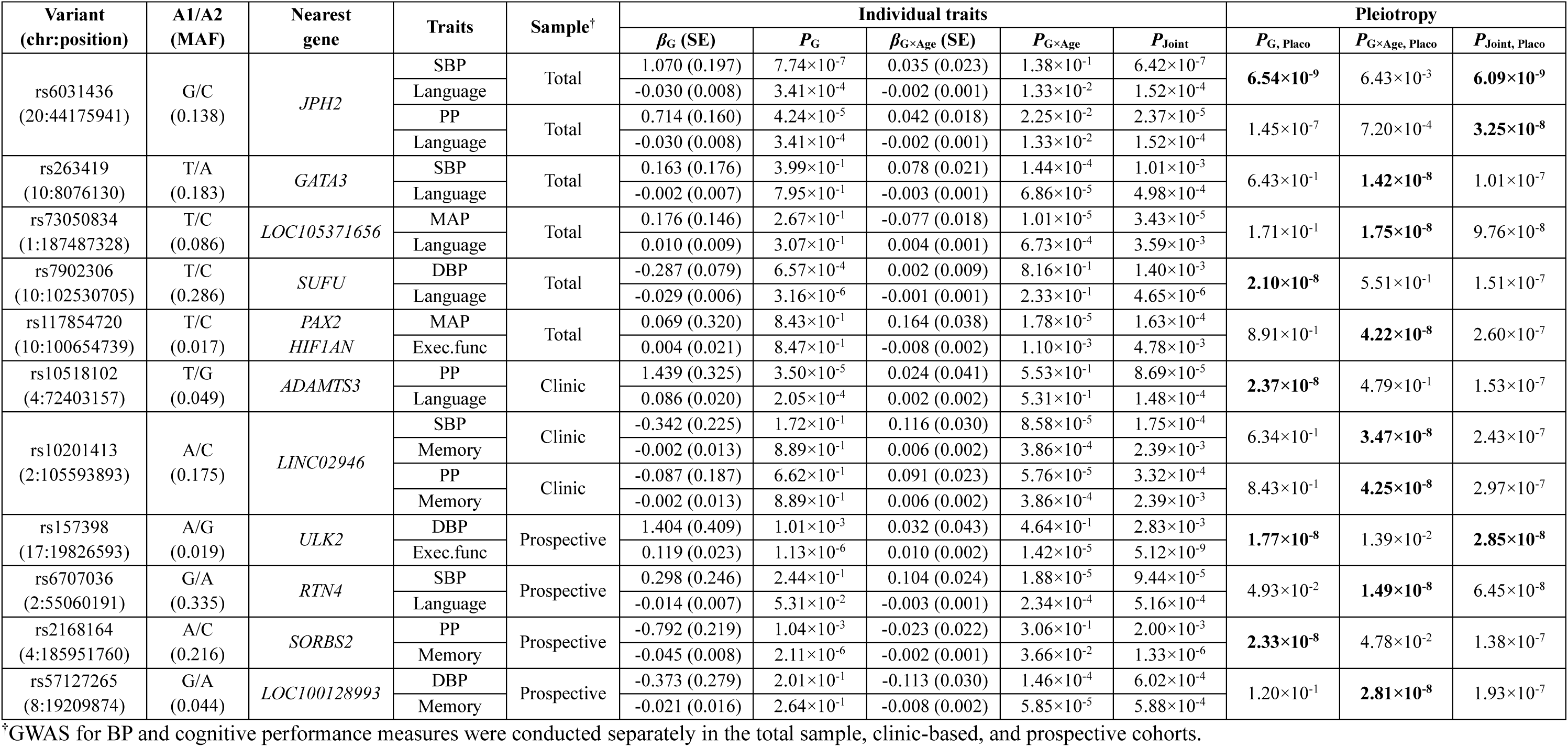
GWS pleiotropy between BP and cognitive performance measures (excluding the *APOE* region)

| Variant<br>(chr:position) | A1/A2<br>(MAF) | Nearest<br>gene | Traits | Sample<sup>†</sup> | Individual traits |  |  |  |  | Pleiotropy |  |  |
| --- | --- | --- | --- | --- | --- | --- | --- | --- | --- | --- | --- | --- |
| | | | | | $\beta_G$ (SE) | $P_G$ | $\beta_{G \times \text{Age}}$ (SE) | $P_{G \times \text{Age}}$ | $P_{\text{Joint}}$ | $P_{G, \text{Placo}}$ | $P_{G \times \text{Age, Placo}}$ | $P_{\text{Joint, Placo}}$ |
| rs6031436<br>(20:44175941) | G/C<br>(0.138) | *JPH2* | SBP | Total | 1.070 (0.197) | $7.74 \times 10^{-7}$ | 0.035 (0.023) | $1.38 \times 10^{-1}$ | $6.42 \times 10^{-7}$ | **$6.54 \times 10^{-9}$** | $6.43 \times 10^{-3}$ | **$6.09 \times 10^{-9}$** |
| | | | Language | | -0.030 (0.008) | $3.41 \times 10^{-4}$ | -0.002 (0.001) | $1.33 \times 10^{-2}$ | $1.52 \times 10^{-4}$ | | | |
| | | | PP | Total | 0.714 (0.160) | $4.24 \times 10^{-5}$ | 0.042 (0.018) | $2.25 \times 10^{-2}$ | $2.37 \times 10^{-5}$ | $1.45 \times 10^{-7}$ | $7.20 \times 10^{-4}$ | **$3.25 \times 10^{-8}$** |
| | | | Language | | -0.030 (0.008) | $3.41 \times 10^{-4}$ | -0.002 (0.001) | $1.33 \times 10^{-2}$ | $1.52 \times 10^{-4}$ | | | |
| rs263419<br>(10:8076130) | T/A<br>(0.183) | *GATA3* | SBP | Total | 0.163 (0.176) | $3.99 \times 10^{-1}$ | 0.078 (0.021) | $1.44 \times 10^{-4}$ | $1.01 \times 10^{-3}$ | $6.43 \times 10^{-1}$ | **$1.42 \times 10^{-8}$** | $1.01 \times 10^{-7}$ |
| | | | Language | | -0.002 (0.007) | $7.95 \times 10^{-1}$ | -0.003 (0.001) | $6.86 \times 10^{-5}$ | $4.98 \times 10^{-4}$ | | | |
| rs73050834<br>(1:187487328) | T/C<br>(0.086) | *LOC105371656* | MAP | Total | 0.176 (0.146) | $2.67 \times 10^{-1}$ | -0.077 (0.018) | $1.01 \times 10^{-5}$ | $3.43 \times 10^{-5}$ | $1.71 \times 10^{-1}$ | **$1.75 \times 10^{-8}$** | $9.76 \times 10^{-8}$ |
| | | | Language | | 0.010 (0.009) | $3.07 \times 10^{-1}$ | 0.004 (0.001) | $6.73 \times 10^{-4}$ | $3.59 \times 10^{-3}$ | | | |
| rs7902306<br>(10:102530705) | T/C<br>(0.286) | *SUFU* | DBP | Total | -0.287 (0.079) | $6.57 \times 10^{-4}$ | 0.002 (0.009) | $8.16 \times 10^{-1}$ | $1.40 \times 10^{-3}$ | **$2.10 \times 10^{-8}$** | $5.51 \times 10^{-1}$ | $1.51 \times 10^{-7}$ |
| | | | Language | | -0.029 (0.006) | $3.16 \times 10^{-6}$ | -0.001 (0.001) | $2.33 \times 10^{-1}$ | $4.65 \times 10^{-6}$ | | | |
| rs117854720<br>(10:100654739) | T/C<br>(0.017) | *PAX2*<br>*HIF1AN* | MAP | Total | 0.069 (0.320) | $8.43 \times 10^{-1}$ | 0.164 (0.038) | $1.78 \times 10^{-5}$ | $1.63 \times 10^{-4}$ | $8.91 \times 10^{-1}$ | **$4.22 \times 10^{-8}$** | $2.60 \times 10^{-7}$ |
| | | | Exec.func | | 0.004 (0.021) | $8.47 \times 10^{-1}$ | -0.008 (0.002) | $1.10 \times 10^{-3}$ | $4.78 \times 10^{-3}$ | | | |
| rs10518102<br>(4:72403157) | T/G<br>(0.049) | *ADAMTS3* | PP | Clinic | 1.439 (0.325) | $3.50 \times 10^{-5}$ | 0.024 (0.041) | $5.53 \times 10^{-1}$ | $8.69 \times 10^{-5}$ | **$2.37 \times 10^{-8}$** | $4.79 \times 10^{-1}$ | $1.53 \times 10^{-7}$ |
| | | | Language | | 0.086 (0.020) | $2.05 \times 10^{-4}$ | 0.002 (0.002) | $5.31 \times 10^{-1}$ | $1.48 \times 10^{-4}$ | | | |
| rs10201413<br>(2:105593893) | A/C<br>(0.175) | *LINC02946* | SBP | Clinic | -0.342 (0.225) | $1.72 \times 10^{-1}$ | 0.116 (0.030) | $8.58 \times 10^{-5}$ | $1.75 \times 10^{-4}$ | $6.34 \times 10^{-1}$ | **$3.47 \times 10^{-8}$** | $2.43 \times 10^{-7}$ |
| | | | Memory | | -0.002 (0.013) | $8.89 \times 10^{-1}$ | 0.006 (0.002) | $3.86 \times 10^{-4}$ | $2.39 \times 10^{-3}$ | | | |
| | | | PP | Clinic | -0.087 (0.187) | $6.62 \times 10^{-1}$ | 0.091 (0.023) | $5.76 \times 10^{-5}$ | $3.32 \times 10^{-4}$ | $8.43 \times 10^{-1}$ | **$4.25 \times 10^{-8}$** | $2.97 \times 10^{-7}$ |
| | | | Memory | | -0.002 (0.013) | $8.89 \times 10^{-1}$ | 0.006 (0.002) | $3.86 \times 10^{-4}$ | $2.39 \times 10^{-3}$ | | | |
| rs157398<br>(17:19826593) | A/G<br>(0.019) | *ULK2* | DBP | Prospective | 1.404 (0.409) | $1.01 \times 10^{-3}$ | 0.032 (0.043) | $4.64 \times 10^{-1}$ | $2.83 \times 10^{-3}$ | **$1.77 \times 10^{-8}$** | $1.39 \times 10^{-2}$ | **$2.85 \times 10^{-8}$** |
| | | | Exec.func | | 0.119 (0.023) | $1.13 \times 10^{-6}$ | 0.010 (0.002) | $1.42 \times 10^{-5}$ | $5.12 \times 10^{-9}$ | | | |
| rs6707036<br>(2:55060191) | G/A<br>(0.335) | *RTN4* | SBP | Prospective | 0.298 (0.246) | $2.44 \times 10^{-1}$ | 0.104 (0.024) | $1.88 \times 10^{-5}$ | $9.44 \times 10^{-5}$ | $4.93 \times 10^{-2}$ | **$1.49 \times 10^{-8}$** | $6.45 \times 10^{-8}$ |
| | | | Language | | -0.014 (0.007) | $5.31 \times 10^{-2}$ | -0.003 (0.001) | $2.34 \times 10^{-4}$ | $5.16 \times 10^{-4}$ | | | |
| rs2168164<br>(4:185951760) | A/C<br>(0.216) | *SORBS2* | PP | Prospective | -0.792 (0.219) | $1.04 \times 10^{-3}$ | -0.023 (0.022) | $3.06 \times 10^{-1}$ | $2.00 \times 10^{-3}$ | **$2.33 \times 10^{-8}$** | $4.78 \times 10^{-2}$ | $1.38 \times 10^{-7}$ |
| | | | Memory | | -0.045 (0.008) | $2.11 \times 10^{-6}$ | -0.002 (0.001) | $3.66 \times 10^{-2}$ | $1.33 \times 10^{-6}$ | | | |
| rs57127265<br>(8:19209874) | G/A<br>(0.044) | *LOC100128993* | DBP | Prospective | -0.373 (0.279) | $2.01 \times 10^{-1}$ | -0.113 (0.030) | $1.46 \times 10^{-4}$ | $6.02 \times 10^{-4}$ | $1.20 \times 10^{-1}$ | **$2.81 \times 10^{-8}$** | $1.93 \times 10^{-7}$ |
| | | | Memory | | -0.021 (0.016) | $2.64 \times 10^{-1}$ | -0.008 (0.002) | $5.85 \times 10^{-5}$ | $5.88 \times 10^{-4}$ | | | |

<sup>†</sup>GWAS for BP and cognitive performance measures were conducted separately in the total sample, clinic-based, and prospective cohorts.

GWS pleiotropy for nine additional loci was identified with the main SNP effect or SNP×age interaction terms (**Tables 2 and S4**). In the total sample, pleiotropy for the SNP×age interaction effect was observed with *GATA3* SNP rs263419 (**Figure 1c**) for SBP paired with language (*P*_G×Age_=1.42×10^-8^), *LOC105371656* SNP rs73050834 (**Figure S6a**) for MAP paired with language (*P*_G×Age_=1.75×10^-8^), and rs117854720 located between *HIF1AN* and *PAX2* (**Figure S6b**) for MAP paired with executive function (*P*_G×Age_=4.22×10^-8^) (**Tables 2 and S4**). The main effect of *SUFU* SNP rs7902306 was associated with the paired outcomes of DBP and language (*P*_G_=2.10×10^-8^), a finding that was supported by several variants in high LD (**Figure 1d**). In the clinic-based cohorts, pleiotropy for the main SNP effect was observed with *ADAMTS3* SNP rs10518102 (**Figure 2b**) for PP paired with language (*P*_G_=2.37×10^-8^), and the SNP×age interaction effect of *LINC02946* SNP rs10201413 was associated with the paired outcomes of SBP and memory (*P*_G×Age_=3.47×10^-8^) and PP and memory (*P*_G×Age_=4.25×10^-8^) (**Figures S6c and S6d**). In the prospective cohorts, pleiotropy for the SNP×age interaction effect was observed with *RTN4* SNP rs6707036 (**Figure 2c**) for SBP paired with language (*P*_G×Age_=1.49×10^-8^) and *LOC100128993* SNP rs57127265 (**Figure S6e**) for DBP paired with memory (*P*_G×Age_=2.81×10^-8^). The main effect of *SORBS2* SNP rs2168164 was also associated with the paired outcomes of PP and memory (*P*_G_=2.33×10^-8^) (**Figure 2d**). MR-Egger analysis revealed that the influence on cognition of five of the 11 GWS pleiotropic loci, including *SUFU* (*P*=4.12×10^-9^), *RTN4* (*P*=5.17×10^-7^), *SORBS2* (*P*=4.58×10^-6^), *ADAMTS3* (*P*=2.20×10^-2^), and *GATA3* (*P*=3.21×10^-2^), was direct rather than through BP (**Table 3**).

**Table 3.** Differentiation of the directional pleiotropy of top-ranked genes from the mediated pleiotropy.

| Gene | Sentinel SNP<br>(chr:position) | Number of<br>LD SNPs<sup>†</sup> | Sample | Traits | *P*-value<sup>‡</sup> |
| --- | --- | --- | --- | --- | --- |
| *JPH2* | rs6031436<br>(20:44175941) | 27 | Total | SBP | 3.75×10<sup>-1</sup> |
|  |  |  |  | Language |  |
|  |  |  |  | PP | 1.76×10<sup>-1</sup> |
|  |  |  |  | Language |  |
| *GATA3* | rs263419<br>(10:8076130) | 35 | Total | SBP | **3.21×10<sup>-2</sup>** |
|  |  |  |  | Language |  |
| *LOC105371656* | rs73050834<br>(1:187487328) | 21 | Total | MAP | 6.91×10<sup>-1</sup> |
|  |  |  |  | Language |  |
| *SUFU* | rs7902306<br>(10:102530705) | 36 | Total | DBP | **4.12×10<sup>-9</sup>** |
|  |  |  |  | Language |  |
| *PAX2*<br>*HIF1AN* | rs117854720<br>(10:100654739) | 18 | Total | MAP | 2.27×10<sup>-1</sup> |
|  |  |  |  | Exec.func |  |
| *ADAMTS3* | rs10518102<br>(4:72403157) | 31 | Clinic | PP | **2.20×10<sup>-2</sup>** |
|  |  |  |  | Language |  |
| *LINC02946* | rs10201413<br>(2:105593893) | 10 | Clinic | SBP | 1.74×10<sup>-1</sup> |
|  |  |  |  | Memory |  |
|  |  |  |  | PP | 4.61×10<sup>-1</sup> |
|  |  |  |  | Memory |  |
| *ULK2* | rs157398<br>(17:19826593) | 20 | Prospective | DBP | 5.18×10<sup>-2</sup> |
|  |  |  |  | Exec.func |  |
| *RTN4* | rs6707036<br>(2:55060191) | 159 | Prospective | SBP | **5.17×10<sup>-7</sup>** |
|  |  |  |  | Language |  |
| *SORBS2* | rs2168164<br>(4:185951760) | 25 | Prospective | PP | **4.58×10<sup>-6</sup>** |
|  |  |  |  | Memory |  |
| *LOC100128993* | rs57127265<br>(8:19209874) | 11 | Prospective | DBP | 6.01×10<sup>-2</sup> |
|  |  |  |  | Memory |  |

<sup>†</sup>MR-Egger analysis included SNPs in LD with a sentinel SNP for each locus. <sup>‡</sup>*P*-value for the MR-Egger intercept test; Genes with a *P*-value less than 0.05 may affect cognition directly or via confounders other than BP (directional pleiotropy). Insignificant genes may affect cognition through BP (mediated pleiotropy).

### Differentially expressed top-ranked pleiotropic genes

Among genes containing or closer than 100 kb to top-ranked GWS pleiotropic SNPs, six genes (*ACTR1A*, *HIF1AN*, *ADAMTS3*, *RTN4*, *SORBS2*, and *SUFU*) were significantly differentially expressed in DLPFC from pathologically confirmed AD cases with and without clinical symptoms prior to death compared to that from cognitively and pathologically normal controls (**Figure 3 and Table S6**). *ACTR1A*, which contains many high LD variants of the top-ranked *SUFU* SNP rs7902306, had significantly lower expression levels (FDR=1.27×10^-4^) in symptomatic AD cases than controls (**Figure 3a**). *HIF1AN*, located near the top-ranked *PAX2* SNP rs117854720, was significantly underexpressed in symptomatic AD cases compared to controls (FDR=4.26×10^-3^) (**Figure 3b**). *ADAMTS3* and *RTN4* had significantly lower expression levels in symptomatic AD cases compared to controls (FDR < 1.31×10^-7^) and resilient AD cases (FDR < 9.78×10^-3^) (**Figures 3c and 3d**). Expression of *SORBS2* was also significantly lower (FDR=1.39×10^-4^) in symptomatic AD cases compared to controls (**Figure 3e**). *SUFU* was significantly overexpressed in symptomatic AD cases compared to controls (FDR=3.91×10^-2^) (**Figure 3f**). None of the top-ranked or suggestive pleiotropic variants were associated with the expression of any of the differentially expressed genes.

**Figure 3.**
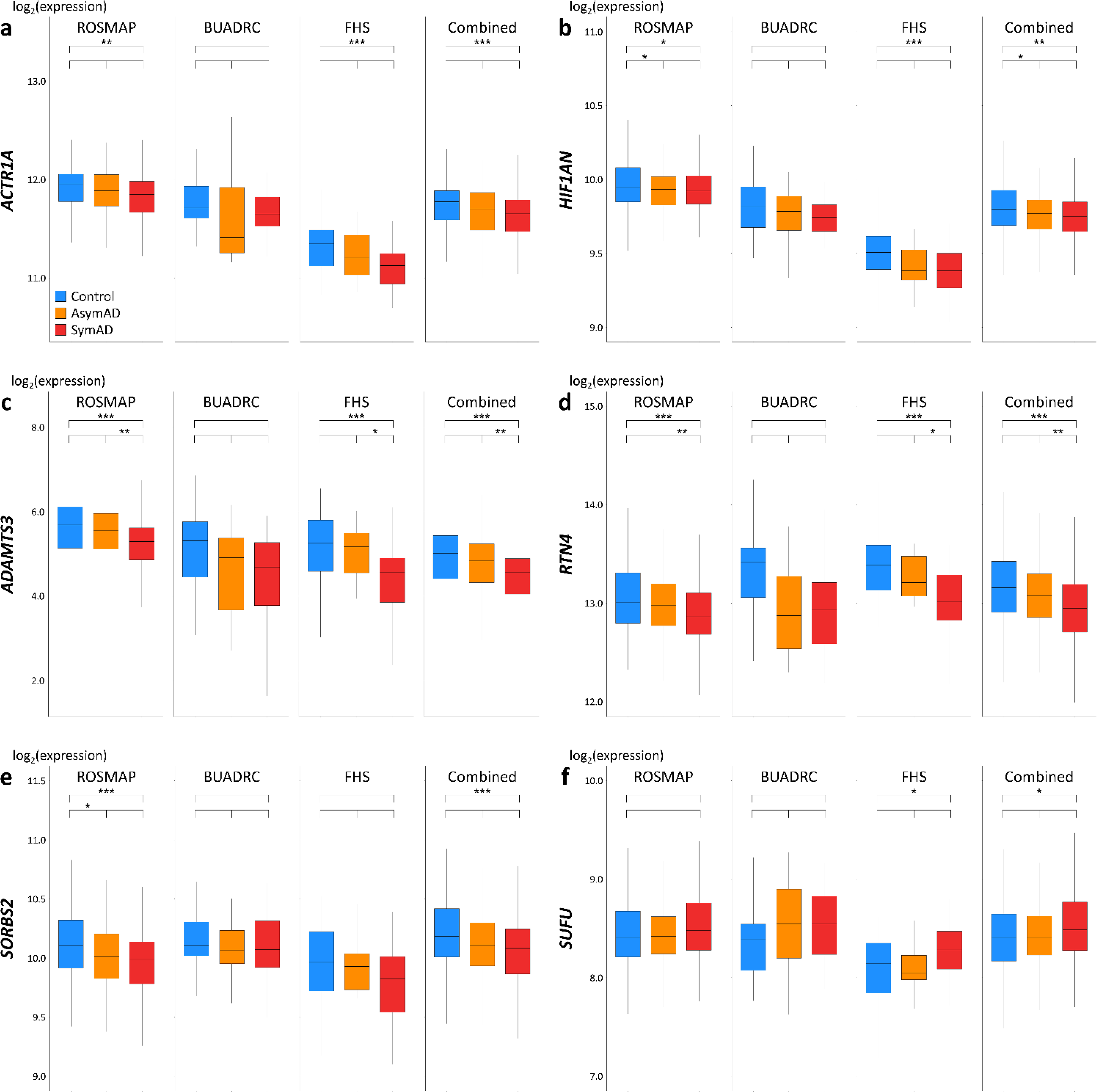
Pleiotropic genes significantly differentially expressed in the DLPFC in pathologically diagnosed AD cases with and without clinical symptoms prior to death and cognitively normal controls. Genes shown are significantly differentially expressed in the DLPFC among pathologically confirmed symptomatic AD (SymAD) cases, asymptomatic AD (AsymAD) cases, and cognitively normal controls. Differential expression levels in the DLPFC were estimated and meta-analyzed using the ROSMAP, BUADRC, and FHS datasets. (a) *ACTR1A*, (b) *HIF1AN*, (c) *ADAMTS3*, (d) *RTN4*, (e) *SORBS2*, and (f) *SUFU*. Units on the Y-axis represent log-transformed (log2) expression of genes. *FDR < 0.05, **FDR < 0.01, ***FDR < 0.001.

### Pathways enriched for top-ranked pleiotropic genes

We identified many canonical pathways significantly enriched for genes containing top-ranked pleiotropic variants (*P* < 1×10^-4^) (**Table S7**), primarily derived from analyses seeded with genes affecting memory paired with SBP or DBP (**Table 4**). Multiple pathways closely connected to G protein-coupled receptors (GPCRs) and their downstream signaling pathways were significantly enriched for genes with pleiotropy between SBP and memory (FDR < 1.20×10^-3^), including orexin signaling, apelin endothelial signaling, P2Y purinergic receptor signaling, protein kinase A (PKA) signaling, γ-aminobutyric acid (GABA)-related (*e*.*g*., GABAergic) receptor signaling, glutaminergic receptor signaling, Gα12/13 signaling, and signaling by rho family GTPases. We also found significant pathways linked to the renin-angiotensin system (RAS) and cardiac hypertrophy (FDR < 1.20×10^-3^), such as the role of nuclear factor of activated T cells in cardiac hypertrophy, renin-angiotensin signaling, cell junction organization, cardiac hypertrophy signaling, and gap junction signaling. Several pathways related to pro-inflammatory responses of microglia and mast cells were significantly enriched (FDR < 8.91×10^-4^), including oxytocin signaling, docosahexaenoic acid signaling, leptin signaling, eicosanoid signaling, relaxin signaling, and interleukin-1 signaling. Other pathways significantly enriched for top-ranked pleiotropic gene were related to insulin resistance (*e*.*g*., apelin pancreas signaling, type II diabetes mellitus signaling, white adipose tissue browning pathway, and insulin receptor signaling) or neuronal development signaling, such as synaptogenesis signaling, gonadotropin-releasing hormone signaling, and endocannabinoid developing neuron pathway.

**Table 4.** Pathways significantly enriched for top-ranked pleiotropic genes (with FDR < 0.001)

| BP trait | Cognitive domain | Ingenuity canonical pathway | Number of seed genes | FDR† |
| --- | --- | --- | --- | --- |
| SBP | Memory | Orexin Signaling Pathway | 299 | 3.16×10<sup>-5</sup> |
|  |  | Role of NFAT in Cardiac Hypertrophy |  | 5.62×10<sup>-5</sup> |
|  |  | Apelin Endothelial Signaling Pathway |  | 1.66×10<sup>-4</sup> |
|  |  | Molecular Mechanisms of Cancer |  | 1.86×10<sup>-4</sup> |
|  |  | Colorectal Cancer Metastasis Signaling |  | 1.86×10<sup>-4</sup> |
|  |  | Renin-Angiotensin Signaling |  | 1.86×10<sup>-4</sup> |
|  |  | Cell junction organization |  | 1.86×10<sup>-4</sup> |
|  |  | Oxytocin Signaling Pathway |  | 1.86×10<sup>-4</sup> |
|  |  | Cardiac Hypertrophy Signaling (Enhanced) |  | 2.19×10<sup>-4</sup> |
|  |  | Docosahexaenoic Acid (DHA) Signaling |  | 2.19×10<sup>-4</sup> |
|  |  | Apelin Pancreas Signaling Pathway |  | 2.19×10<sup>-4</sup> |
|  |  | P2Y Purinergic Receptor Signaling Pathway |  | 2.19×10<sup>-4</sup> |
|  |  | Leptin Signaling in Obesity |  | 3.16×10<sup>-4</sup> |
|  |  | Protein Kinase A Signaling |  | 3.16×10<sup>-4</sup> |
|  |  | Synaptogenesis Signaling Pathway |  | 3.31×10<sup>-4</sup> |
|  |  | Endocannabinoid Cancer Inhibition Pathway |  | 3.72×10<sup>-4</sup> |
|  |  | GNRH Signaling |  | 4.27×10<sup>-4</sup> |
|  |  | Eicosanoid Signaling |  | 4.27×10<sup>-4</sup> |
|  |  | Type II Diabetes Mellitus Signaling |  | 4.37×10<sup>-4</sup> |
|  |  | Relaxin Signaling |  | 4.57×10<sup>-4</sup> |
|  |  | Endocannabinoid Developing Neuron Pathway |  | 7.24×10<sup>-4</sup> |
|  |  | IL-1 Signaling |  | 8.91×10<sup>-4</sup> |
|  |  | Gα12/13 Signaling |  | 9.33×10<sup>-4</sup> |
| DBP | Memory | Insulin Receptor Signaling | 281 | 3.72×10<sup>-4</sup> |
|  |  | Synaptogenesis Signaling Pathway |  | 3.72×10<sup>-4</sup> |

†Benjamini-Hochberg procedure was applied in multiple hypothesis testing to control a false discovery rate.

## Discussion

### Variants in multiple genes contribute to both BP and cognitive decline

Genome-wide pleiotropy analyses for four BP measures paired with performance scores for three cognitive domains identified GWS pleiotropy with *APOE* and 11 novel loci. Among the novel loci, the effect on cognition of five (*SUFU*, *RTN4*, *SORBS2*, *ADAMTS3*, and *GATA3*) was direct rather than through BP, and six genes (*ACTR1A*, *HIF1AN*, *ADAMTS3*, *RTN4*, *SORBS2*, and *SUFU*) containing or closest to the top-ranked GWS pleiotropic SNPs were significantly differentially expressed in DLPFC from pathologically confirmed AD cases with antemortem AD clinical symptoms compared to that from cognitively normal controls and cognitively resilient pathologically confirmed AD cases. We also identified many pathways implicated in high BP and AD/ADRD, which were significantly enriched for genes seeded with top-ranked pleiotropic variants. Our findings were based on time-varying analyses considering repeated measures of BP and cognitive domain scores and their changes over time. Using a model with terms for the SNP main effect and SNP interaction with age, we investigated pleiotropic associations in the total, clinic-based, and prospective cohort samples separately, which enabled us to include data collected at multiple time points and find associations that are age-dependent.

### Roles of pleiotropic loci in AD/ADRD

We identified four loci functionally relevant to processes implicated in AD/ADRD. *ADAMTS3* encodes a major proteolytic enzyme that breaks down and inactivates reelin in the cerebral cortex and hippocampus [70], and reelin plays a crucial role in resilience to AD [71]. Increased *RTN4* expression inhibits the cleavage of amyloid precursor protein (APP) by blocking the access of β-site APP cleaving enzyme 1 (BACE1) to APP and thus decreases Aβ production [72]. Nogo-A, also known as RTN4-A, promotes Aβ secretion via the Nogo-66 receptor/ROCK-dependent BACE1 pathway, leading to the onset and development of AD/ADRD [73]. Nogo-B, another member of the RTN4 family, regulates endothelial sphingolipid biosynthesis and promotes endothelial dysfunction and high BP [74]. *ULK2*-dependent mitophagy activation is required for synaptic toxicity in cortical neurons and hippocampal CA1 neurons, triggered by AMP-activated protein kinase overactivated by Aβ42 oligomers [75]. We also identified a GWS association of DBP with *ULK4*, another member of the *ULK* family. The fusion of a duplication of several *ULK4* exons with a partial duplication of neighboring gene *CHRNA7* [76] forms *CHRFAM7A*, a negative regulator of alpha7 nicotinic acetylcholine receptor (α7nAChR) which has a high affinity for Aβ and plays a role in Aβ-induced neuroinflammation [77]. *CHRFAM7A* reduces Aβ uptake via α7nAChR and acts as an immune switch that shifts the α7nAChR from anti-inflammatory to pro-inflammatory, particularly in microglia-like cells [77]. *HIF1AN* encodes an inhibitor of hypoxia-inducible factor-1α (HIF-1α) whose activation leads to excessive Aβ deposition and tau hyperphosphorylation by abnormally cleaving APP and inhibiting Aβ degradation [78]. HIF-1α also controls BP homeostasis, and the loss of HIF-1α in vascular smooth muscle (VSM) cells leads to hypertension in vivo [79].

*GATA3* and *SUFU* are functionally relevant to the immune system (*e*.*g*., pro-inflammatory signaling activation and inflammatory responses), which may enhance neuroinflammatory responses. *GATA3* has been associated with female-specific cognitive resilience to AD pathology [80] and encodes a transcription factor that controls T helper 2 cells which produce Aβ auto-antibodies, alleviate Aβ deposition, and are therefore protective against AD [81]. *GATA3* activates Tie2 promotor and regulates angiopoietin-1/Tie2 signaling, inhibition of which may result in endothelial dysfunction [82]. *SUFU* encodes a negative regulator of the sonic hedgehog signaling pathway modulating immune responses [83]. *ACTR1A* located about 100 bp upstream of *SUFU* is linked to retrograde axonal transport [84], dysregulation of which is known to be an early event in AD/ADRD [85]. *SUFU* and *ACTR1A* also have been linked to BP in a large trans-ethnic GWAS [13]. *JPH2* plays a critical role in maintaining the effective flux of calcium ions [86], dysregulation of which is related to neuroinflammatory signaling in neurons [87], microglia [88], and astrocytes [89]. Downregulated *JPH2* expression is also linked to hypertrophic cardiomyopathy [90] and increases the risk of AD/ADRD by about 50% [91]. *LINC02946* encodes a long intergenic non-protein coding RNA that is not expressed in brain, however it is located 150 kb upstream of the most proximate protein-coding gene, *NCK2*, a recently identified AD GWAS locus [92]. Roles of *SORBS2*, *LOC105371656*, and *LOC100128993* in mechanisms of AD/ADRD or neuroinflammation are unclear at this time.

### Links of abnormal BP and AD to endothelial dysfunction and neuroinflammation

Analyses of biological pathways and gene networks based on top-ranked genes with pleiotropic effects on BP and cognitive measures identified significant pathways involved in GPCR signaling, RAS, and pro-inflammatory responses. GPCRs have a key role in regulating BACE1 and are involved in the pathogenesis of AD/ADRD [93]. Multiple studies highlighted the vulnerability of GABAergic system in the cerebrospinal fluid of AD patients [94] and the role of PKA dysregulation in AD pathogenesis [95]. GPCRs expressed in endothelial and VSM cells also contribute to maintaining vascular homeostasis and play a critical role in BP regulation [96]. RAS contributes to endothelial dysfunction that exacerbates hypertensive conditions and leads to neuroinflammatory responses [22, 97, 98]. Pro-inflammatory responses of microglia may trigger neuroinflammation and promote AD/ADRD pathogenesis [99, 100], and pro-inflammatory cytokines accumulated in vessels and kidneys induce vascular and renal damage, promoting a progressive increase in BP [101]. Interestingly, most significant pathways seeded with top-ranked pleiotropic genes were implicated in the memory domain, which is reasonable because memory impairment is one of the most prominent cognitive features in AD/ADRD.

The potential mechanistic links between late-life BP changes and AD are evident in many studies and imply the existence of shared genetics between BP and cognitive performance, also tangled with immune responses. A late-life PP increase is mainly caused by arterial stiffness [102] and is associated with endothelial dysfunction [103]. Under normal conditions, endothelium balances the release of vasoconstrictors and vasodilators to regulate vascular tone and blood flow (BF) [104] in response to shear stress (*i*.*e*., the BF-mediated frictional force exerted on the vessel wall [105]). Under chronic hypertensive conditions, however, the release of a crucial vasodilator nitric oxide (NO) is depressed [106], and endothelium loses the ability to maintain vascular homeostasis. This endothelial dysfunction is mainly due to activated NADH/NADPH oxidase and the RAS, leading to arterial superoxide overproduction and endothelium-derived NO reduction [97]. Moreover, excessive shear stress or high BP-induced endothelial dysfunction exacerbates hypertensive conditions and accelerates the infiltration of peripheral immune cells, originating outside the central nervous system, into the brain by disturbing the blood-brain barrier and activating pro-inflammatory signaling [22, 98], which leads to neuroinflammatory responses implicated in AD [107, 108] and results in the accumulation of neurotoxic Aβ in the brain [109].

### Pleiotropic loci may contribute to cognitive resilience and inflammation

Expression of five genes (*ACTR1A*, *HIF1AN*, *ADAMTS3*, *RTN4*, and *SORBS2*) containing GWS pleiotropic SNPs was highest in controls, intermediate in cognitively resilient AD cases, and lowest in symptomatic AD cases, suggesting that they may impact the development of AD pathology rather than cognitive impairment. We also found significantly different expressions of *ADAMTS3* and *RTN4* between AD cases with and without clinical symptoms, suggesting their involvement in cognitive resilience, noting that *ADAMTS3* was previously linked to *RELN* which has been implicated in resilience to AD [70, 71].

### Effects of some pleiotropic loci are age-dependent

We applied models with terms for the main SNP and SNP×age interaction effects and jointly tested them to increase the power to detect genetic associations and pleiotropy between BP and cognitive performance measures. However, the interpretation of results emerging from the joint test combining the main SNP and SNP×age interaction effects is not straightforward. Among GWS loci showing pleiotropy between BP and cognitive measures, five loci (*JPH2*, *ULK2*, *SUFU*, *ADAMTS3*, and *SORBS2*) had GWS main SNP effects, suggesting these loci may affect mechanisms that influence BP and cognitive performance rather than changes in these measures over time. In contrast, other loci (*GATA3*, *LOC105371656*, *PAX2*, *LINC02946*, *RTN4*, and *LOC100128993*) showing significant SNP×age interaction effects may affect changes in BP or cognitive measures over time. Considering there were no significant SNP×age interactions in the GWAS of individual BP traits, pairing BP (which typically increases with age) with cognitive performance (which typically decreases with age) may have afforded greater power to detect loci whose effects are age-dependent. Associations with *ADAMTS3* and *LINC02946* were observed only in the clinic-based cohorts in which the onset of AD symptoms on average occurred at a younger age than in the prospective cohorts. Conversely, associations with *ULK2*, *RTN4*, *SORBS2*, and *LOC100128993* were observed only in the prospective cohorts, implying these loci influence AD onset at a later age or may be associated with age-related cognitive decline that is not specific to AD/ADRD processes.

### Study limitations

Our study has several limitations. Except for FHS participants, individuals included in this study were much older at the time of enrollment than those in most BP GWAS. Hence, associations with loci whose effects on BP start in young or middle adulthood may have been missed. Despite including several large longitudinal cohorts, the sample was considerably smaller compared to samples included in BP or AD GWAS, and thus the power for testing the main SNP and SNP×age interaction effects were reduced. Also, a large portion of the participants were examined only once (FHS: 28.6%, ACT: 12.4%, ROSMAP: 7.4%, NACC: 19.3%, and ADNI: 13.0%), and those individuals did not contribute to analyses of changes over time. Furthermore, the interpretation of some of our findings is complicated because they may be implicate multiple mechanisms affecting BP and cognitive performance, including their changes over time. Another concern is the lack of correction for conducting 36 genome-wide pleiotropy analyses (*i*.*e*., 12 pairs of BP and cognitive measures in the total, clinic-based, and prospective cohort samples). Correction for 36 analyses would elevate the GWS threshold to *P* < 1.39×10^-9^, but this corrected threshold may be too conservative because there are strong correlations among the three cognitive domain scores, among the four BP measures, and between the separately ascertained cohorts and total sample. Finally, our findings are specific to non-Hispanic white participants and should be replicated in other population groups to generalize them more broadly.

## Conclusions

Our findings provide additional insight into the underlying mechanisms of hypertension and AD/ADRD, particularly those involving neuroinflammation. Ongoing efforts to harmonize BP and cognitive performance measures across several cohorts would likely yield additional datasets for discovering new, replicating, and generalizing associations with loci influencing measures of both BP and cognitive performance. Further studies are needed to identify mechanisms underlying pleiotropic associations.

## Data Availability

All data produced in the present study are available upon reasonable request to the authors.

## Abbreviations

α7nAChR: alpha7 nicotinic acetylcholine receptor
Aβ: amyloid-β
ACT: Adult Changes in Thought
AD: Alzheimer’s disease
ADNI: Alzheimer’s Disease Neuroimaging Initiative
ADRD: Alzheimer’s disease related dementias
APP: amyloid precursor protein
BACE1: β-site APP cleaving enzyme 1
BF: blood flow
BMI: body mass index
BP: blood pressure
BUADRC: Boston University Alzheimer’s Disease Research Center
DBP: diastolic blood pressure
DLPFC: dorsolateral prefrontal cortex
FDR: false discovery rate
FHS: Framingham Heart Study
GABA: γ-aminobutyric acid
GPCR: G protein-coupled receptor
GRCh38: Genome Research Consortium human build 38
GRM: genetic relationship matrix
GWAS: genome-wide association study
GWS: genome-wide significant
HIF-1α: hypoxia-inducible factor-1α
LD: linkage disequilibrium
MAF: minor allele frequency
MAP: mean arterial pressure
MCI: mild cognitive impairment
MMSE: Mini-Mental State Examination
MR: Mendelian randomization
NACC: National Alzheimer’s Coordinating Center
NIA: National Institute on Aging
NO: nitric oxide
NP: neuropsychological
PC: principal component
PKA: protein kinase A
PP: pulse pressure
QC: quality control
RAS: renin-angiotensin system
ROSMAP: Religious Orders Study/Rush Memory and Aging Project
SBP: systolic blood pressure
SE: standard error
SNP: single nucleotide polymorphism
TOPMed: Trans-Omics for Precision Medicine
VSM: vascular smooth muscle

## Acknowledgements

Biological samples and associated phenotypic data used in primary data analyses were stored at study investigator institutions and at the National Cell Repository for Alzheimer’s Disease (NCRAD, U24-AG021886) at Indiana University funded by the National Institute on Aging (NIA). Associated phenotypic data used in primary and secondary data analyses were provided by the study investigators, the NIA funded Alzheimer’s Disease Centers (ADCs), the National Alzheimer’s Coordinating Center (NACC, U01-AG016976), the National Institute on Aging Genetics of Alzheimer’s Disease Data Storage Site (NIAGADS, U24-AG041689) at the University of Pennsylvania, funded by NIA, and the Religious Orders Study/Rush Memory and Aging Project (ROSMAP, P30-AG10161, P30-AG72975, R01-AG15819, R01-AG17917, U01-AG46152, and U01-AG61356). Phenotypic data were harmonized by the Alzheimer’s Disease Sequencing Project Phenotype Harmonization Consortium (ADSP-PHC, U24-AG074855, U01-AG068057 and R01-AG059716), funded by NIA. This research was supported in part by the Intramural Research Program of the National Institutes of Health (NIH), National Library of Medicine. Contributors to the genetic and phenotypic data included the study investigators on projects that were individually funded by NIA and other NIH institutes, and by private United States organizations, foreign governmental organizations, or nongovernmental organizations. We also acknowledge the following investigators who assembled and characterized participants of cohorts included in this study.

Adult Changes in Thought: James D. Bowen, Paul K. Crane, Gail P. Jarvik, C. Dirk Keene, Eric B. Larson, W. William Lee, Wayne C. McCormick, Susan M. McCurry, Shubhabrata Mukherjee, Katie Rose Richmire

Chicago Health and Aging Project: Philip L. De Jager, Denis A. Evans.

Estudio Familiar de la Influencia Genetica en Alzheimer: Sandra Barral, Rafael Lantigua, Richard Mayeux, Martin Medrano, Dolly Reyes-Dumeyer, Badri Vardarajan.

Framingham Heart Study: Ting Fang Alvin Ang, Hugo J. Aparicio, Rhoda Au, Sanford Auerbach, Alexa S. Beiser, Anita DeStefano, Sherral Devine, Lindsay A. Farrer, Jesse Mez, Jose Raphael Romero, Sudha Seshadri.

Genetic Differences: Duane Beekly, James Bowen, Walter A. Kukull, Eric B. Larson, Wayne McCormick, Gerard D. Schellenberg, Linda Teri.

Mayo Clinic: Minerva M. Carrasquillo, Dennis W. Dickson, Nilufer Ertekin-Taner, Neill R. Graff-Radford, Joseph E. Parisi, Ronald C. Petersen, Steven G. Younkin.

Mayo PD: Gary W. Beecham, Dennis W. Dickson, Ranjan Duara, Nilufer Ertekin-Taner, Tatiana M. Foroud, Neill R. Graff-Radford, Richard B. Lipton, Joseph E. Parisi, Ronald C. Petersen, Bill Scott, Jeffery M. Vance.

Memory and Aging Project: David A. Bennett, Philip L. De Jager.

Multi-Institutional Research in Alzheimer’s Genetic Epidemiology Study: Sanford Auerbach, Helan Chui, Jaeyoon Chung, L. Adrienne Cupples, Charles DeCarli, Ranjan Duara, Martin Farlow, Lindsay A. Farrer, Robert Friedland, Rodney C.P. Go, Robert C. Green, Patrick Griffith, John Growdon, Gyungah R. Jun, Walter Kukull, Alexander Kurz, Mark Logue, Kathryn L. Lunetta, Thomas Obisesan, Helen Petrovitch, Marwan Sabbagh, A. Dessa Sadovnick, Magda Tsolaki.

National Cell Repository for Alzheimer’s Disease: Kelley M. Faber, Tatiana M. Foroud.

National Institute on Aging (NIA) Late Onset Alzheimer’s Disease Family Study: David A. Bennett, Sarah Bertelsen, Thomas D. Bird, Bradley F. Boeve, Carlos Cruchaga, Kelley Faber, Martin Farlow, Tatiana M. Foroud, Alison M. Goate, Neill R. Graff-Radford, Richard Mayeux, Ruth Ottman, Dolly Reyes-Dumeyer, Roger Rosenberg, Daniel Schaid, Robert A. Sweet, Giuseppe Tosto, Debby Tsuang, Badri Vardarajan.

NIA Alzheimer Disease Centers: Erin Abner, Marilyn S. Albert, Roger L. Albin, Liana G. Apostolova, Sanjay Asthana, Craig S. Atwood, Lisa L. Barnes, Thomas G. Beach, David A. Bennett, Eileen H. Bigio, Thomas D. Bird, Deborah Blacker, Adam Boxer, James B. Brewer, James R. Burke, Jeffrey M. Burns, Joseph D. Buxbaum, Nigel J. Cairns, Chuanhai Cao, Cynthia M. Carlsson, Richard J. Caselli, Helena C. Chui, Carlos Cruchaga, Mony de Leon, Charles DeCarli, Malcolm Dick, Dennis W. Dickson, Nilufer Ertekin-Taner, David W. Fardo, Martin R. Farlow, Lindsay A. Farrer, Steven Ferris, Tatiana M. Foroud, Matthew P. Frosch, Douglas R. Galasko, Marla Gearing, David S. Geldmacher, Daniel H. Geschwind, Bernardino Ghetti, Carey Gleason, Alison M. Goate, Teresa Gomez-Isla, Thomas Grabowski, Neill R. Graff-Radford, John H. Growdon, Lawrence S. Honig, Ryan M. Huebinger, Matthew J. Huentelman, Christine M. Hulette, Bradley T. Hyman, Suman Jayadev, Lee-Way Jin, Sterling Johnson, M. Ilyas Kamboh, Anna Karydas, Jeffrey A. Kaye, C. Dirk Keene, Ronald Kim, Neil W. Kowall, Joel H. Kramer, Frank M. LaFerla, James J. Lah, Allan I. Levey, Ge Li, Andrew P. Lieberman, Oscar L. Lopez, Constantine G. Lyketsos, Daniel C. Marson, Ann C. McKee, Marsel Mesulam, Jesse Mez, Bruce L. Miller, Carol A. Miller, Abhay Moghekar, John C. Morris, John M. Olichney, Joseph E. Parisi, Henry L. Paulson, Elaine Peskind, Ronald C. Petersen, Aimee Pierce, Wayne W. Poon, Luigi Puglielli, Joseph F. Quinn, Ashok Raj, Murray Raskind, Eric M. Reiman, Barry Reisberg, Robert A. Rissman, Erik D. Roberson, Howard J. Rosen, Roger N. Rosenberg, Martin Sadowski, Mark A. Sager, David P. Salmon, Mary Sano, Andrew J. Saykin, Julie A. Schneider, Lon S. Schneider, William W. Seeley, Scott Small, Amanda G. Smith, Robert A. Stern, Russell H. Swerdlow, Rudolph E. Tanzi, Sarah E. Tomaszewski Farias, John Q. Trojanowski, Juan C. Troncoso, Debby W. Tsuang, Vivianna M. Van Deerlin, Linda J. Van Eldik, Harry V. Vinters, Jean Paul Vonsattel, Jen Chyong Wang, Sandra Weintraub, Kathleen A. Welsh-Bohmer, Shawn Westaway, Thomas S. Wingo, Thomas Wisniewski, David A. Wolk, Randall L. Woltjer, Steven G. Younkin, Lei Yu, Chang-En Yu.

Religious Orders Study: David A. Bennett, Philip L. De Jager.

Texas Alzheimer’s Research and Care Consortium: Perrie Adams, Alyssa Aguirre, Lisa Alvarez, Gayle Ayres, Robert C. Barber, John Bertelson, Sarah Brisebois, Scott Chasse, Munro Culum, Eveleen Darby, John C. DeToledo, Thomas J. Fairchild, James R. Hall, John Hart, Michelle Hernandez, Ryan Huebinger, Leigh Johnson, Kim Johnson, Aisha Khaleeq, Janice Knebl, Laura J. Lacritz, Douglas Mains, Paul Massman, Trung Nguyen, Sid O’Bryant, Marcia Ory, Raymond Palmer, Valory Pavlik, David Paydarfar, Victoria Perez, Marsha Polk, Mary Quiceno, Joan S. Reisch, Monica Rodriguear, Roger Rosenberg, Donald R. Royall, Janet Smith, Alan Stevens, Jeffrey L. Tilson, April Wiechmann, Kirk C. Wilhelmsen, Benjamin Williams, Henrick Wilms, Martin Woon.

University of Miami: Larry D. Adams, Gary W. Beecham, Regina M. Carney, Katrina Celis, Michael L. Cuccaro, Kara L. Hamilton-Nelson, James Jaworski, Brian W. Kunkle, Eden R. Martin, Margaret A. Pericak-Vance, Farid Rajabli, Michael Schmidt, Jeffery M Vance.

University of Toronto: Ekaterina Rogaeva, Peter St. George-Hyslop.

University of Washington Families: Thomas D. Bird, Olena Korvatska, Wendy Raskind, Chang-En Yu.

Vanderbilt University: John H. Dougherty, Harry E. Gwirtsman, Jonathan L. Haines, Angela Jefferson.

Washington Heights-Inwood Columbia Aging Project: Adam Brickman, Rafael Lantigua, Jennifer Manly, Richard Mayeux, Christiane Reitz, Nicole Schupf, Yaakov Stern, Giuseppe Tosto, Badri Vardarajan.

