## Supplemental Figures for "Genome-wide pleiotropy analysis of longitudinal blood pressure and harmonized cognitive performance measures"

**Figure S1. Schemes for combining results from GWAS and pleiotropy analyses across datasets (total sample)**

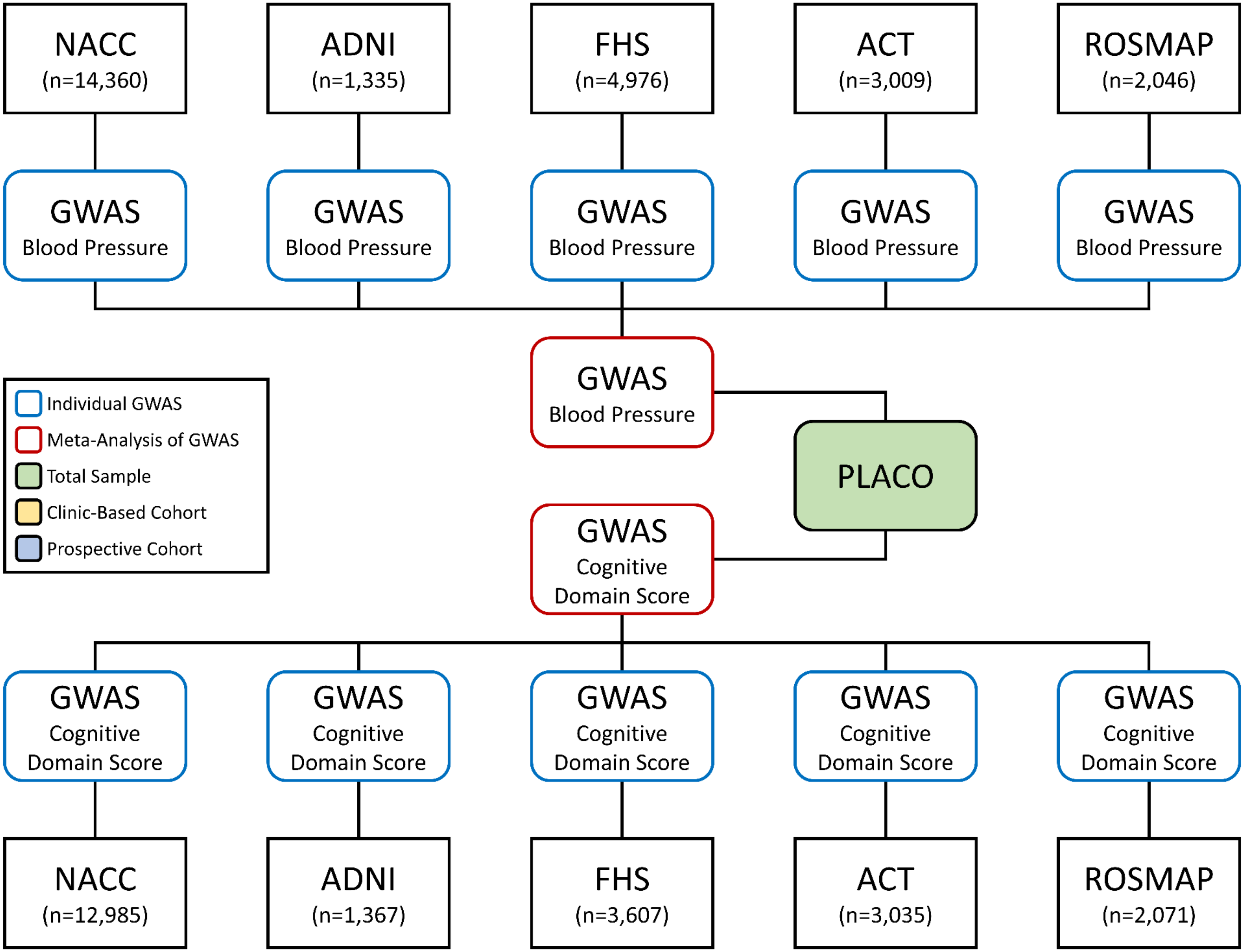

**Figure S1. Continued (clinic-based and prospective cohorts)**

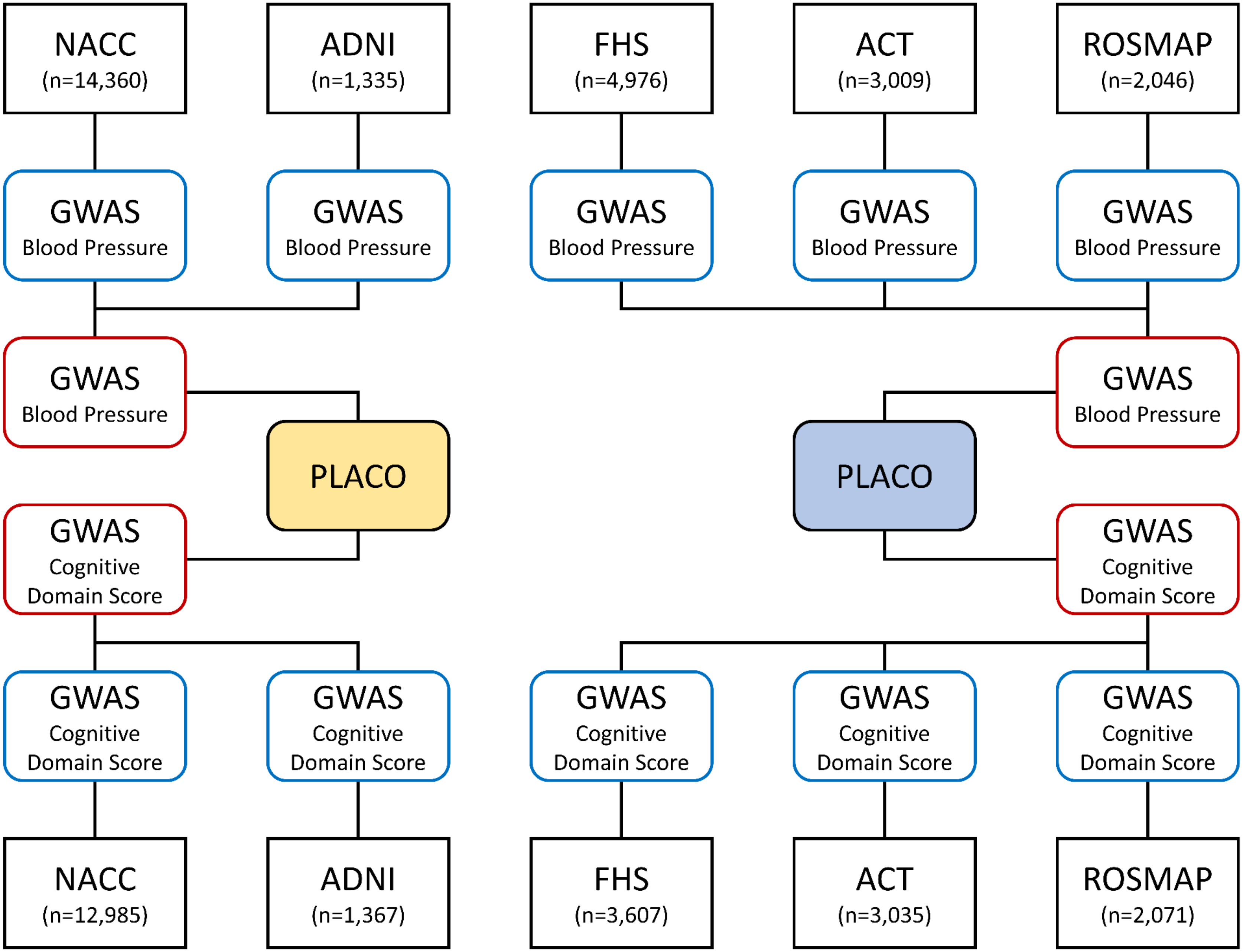

**Figure S2. Phenotypic and genetic correlations among BP and cognitive performance measures (phenotypic correlations)**

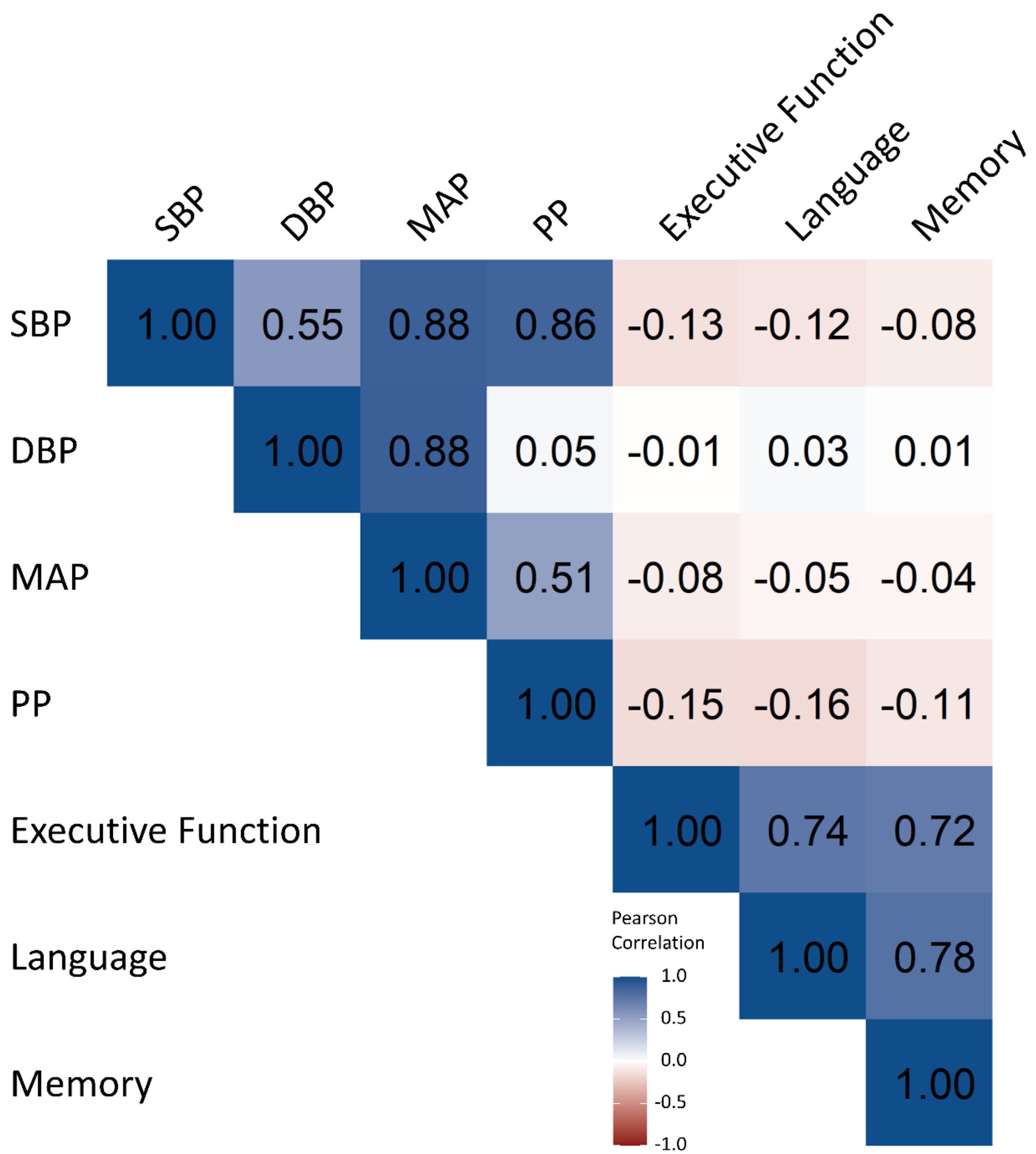

**Figure S2. Continued (genetic correlations)**

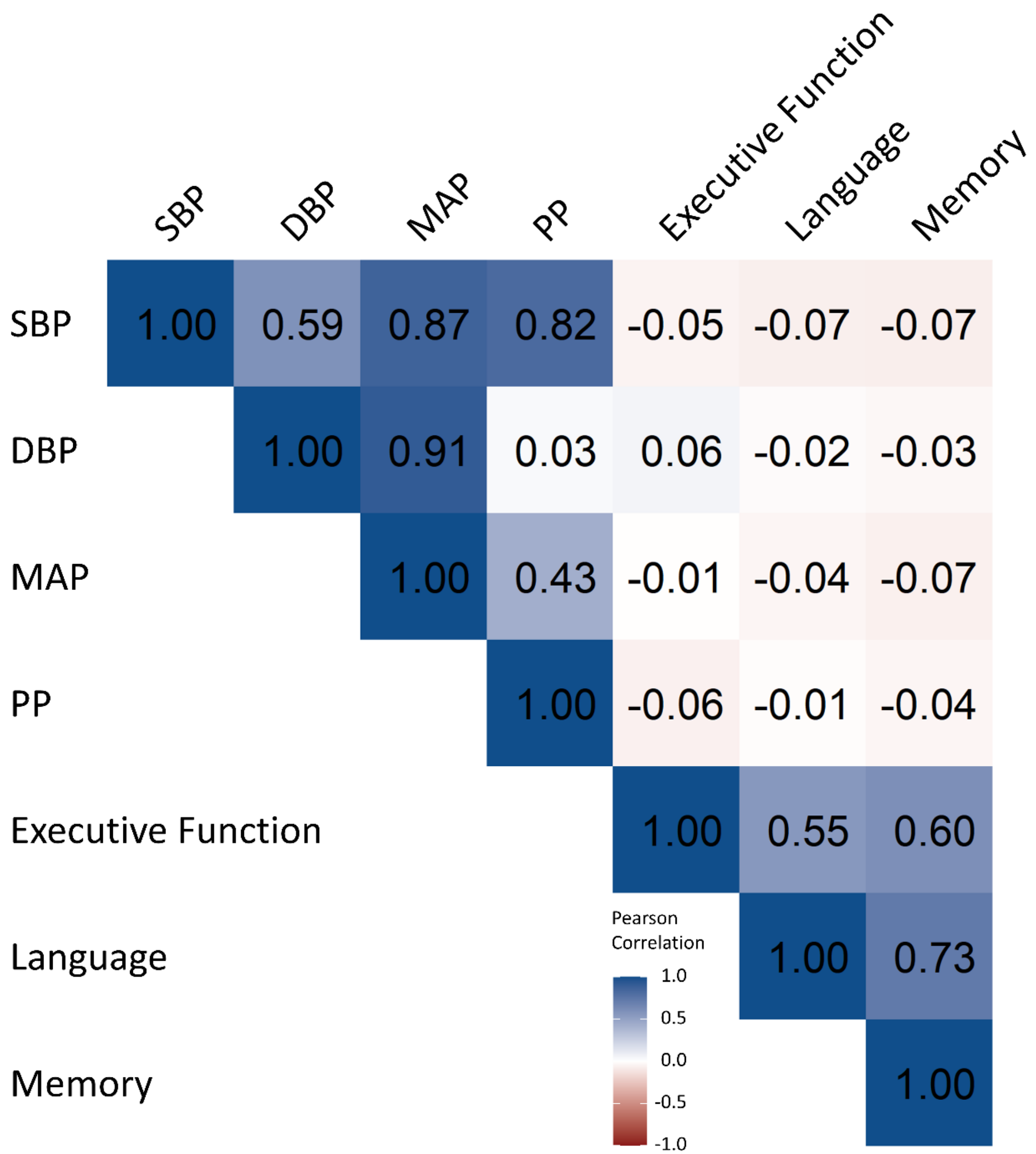

### Figure S3. Manhattan and QQ plots for individual BP GWAS (total sample)

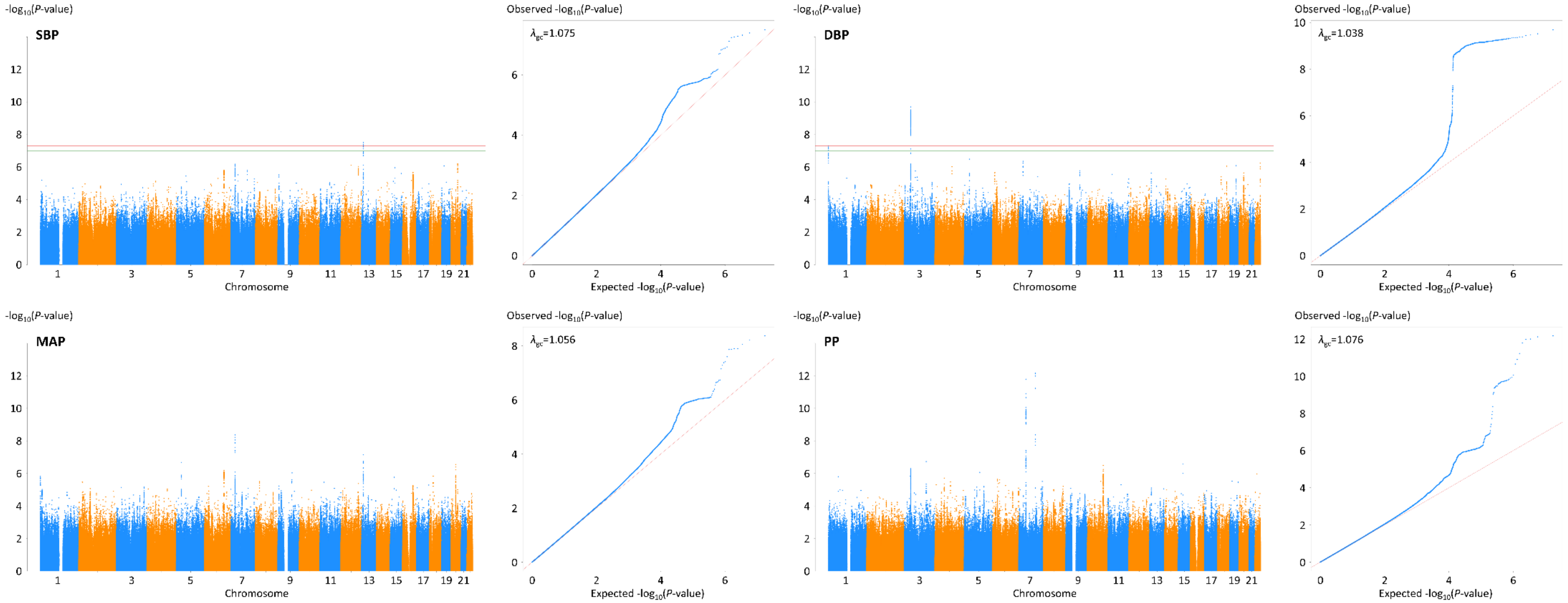

### Figure S3. Continued (clinic-based cohorts)

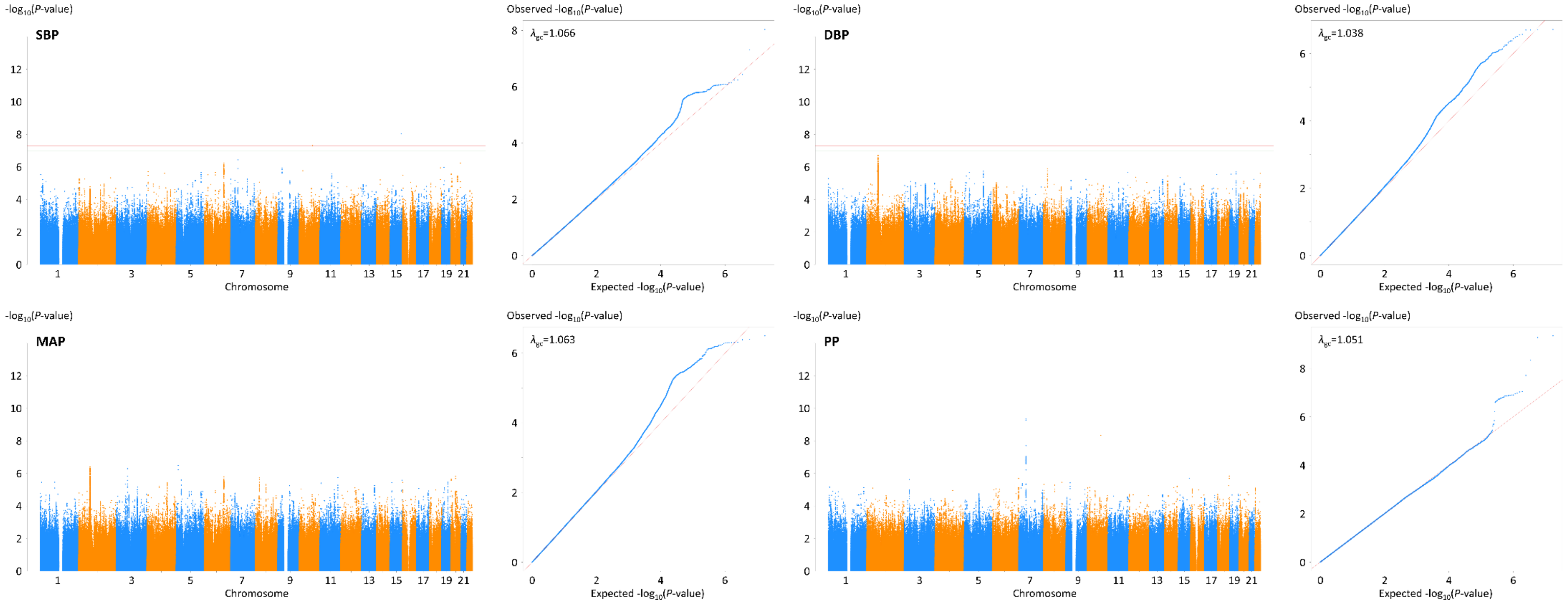

### Figure S3. Continued (prospective cohorts)

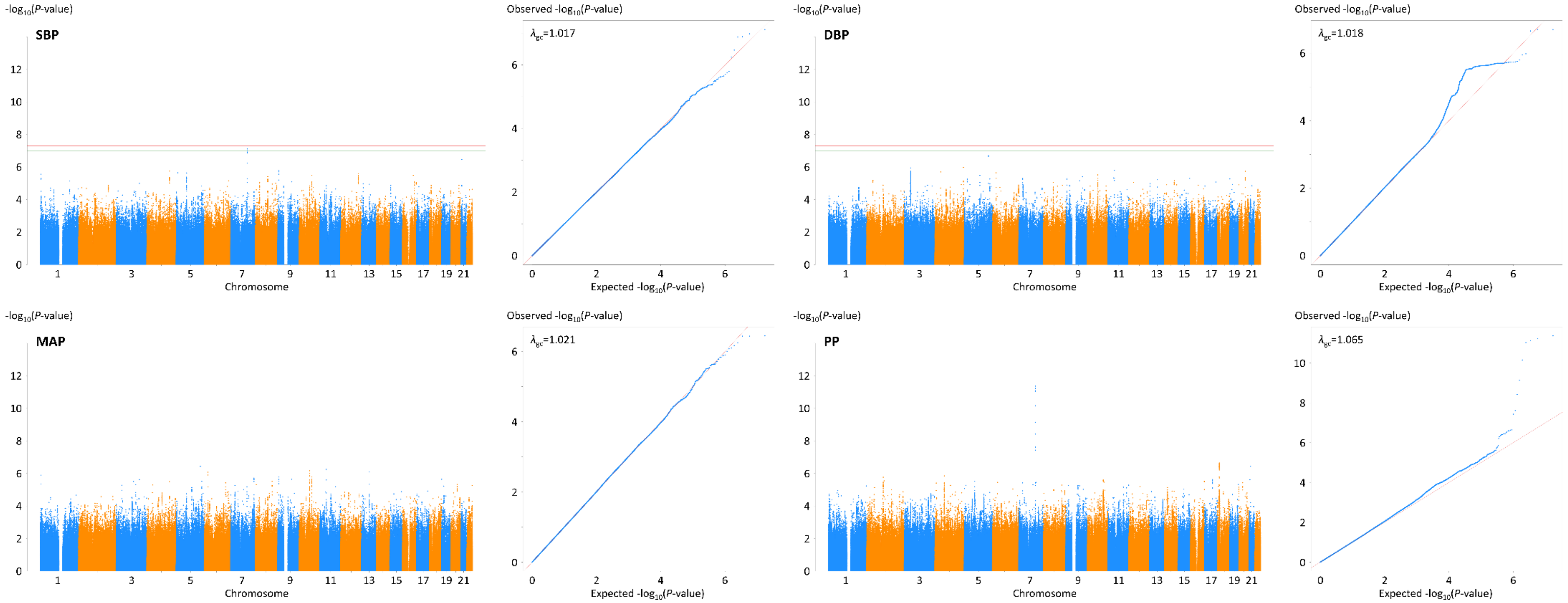

**Figure S4. LocusZoom plots showing SNP associations in top-ranked BP loci.** (a) rs6490411 and *SLC7A1* variants associated with SBP in the total sample, (b) rs57653201 and *ULK4* variants associated with DBP in the total sample, (c) rs60772526 and *HOTTIP* variants associated with MAP in the total sample, and (d) rs12705390 and *PIK3CG* variants associated with PP in the total sample. A purple diamond indicates the top-ranked SNP at each locus. SNPs are color-coded according to their LD ($r^2$) with the top-ranked SNP in the region. Horizontal dotted lines represent the GWS threshold of $P$=5×10$^{-8}$. Vertical blue lines indicate locations of the high recombination rate among SNPs at the chromosomal position. Approximate location, transcription direction, and coding portions (exons represented by vertical bars) of genes are shown below the diagram. Mb=megabase, cM=centimorgan.

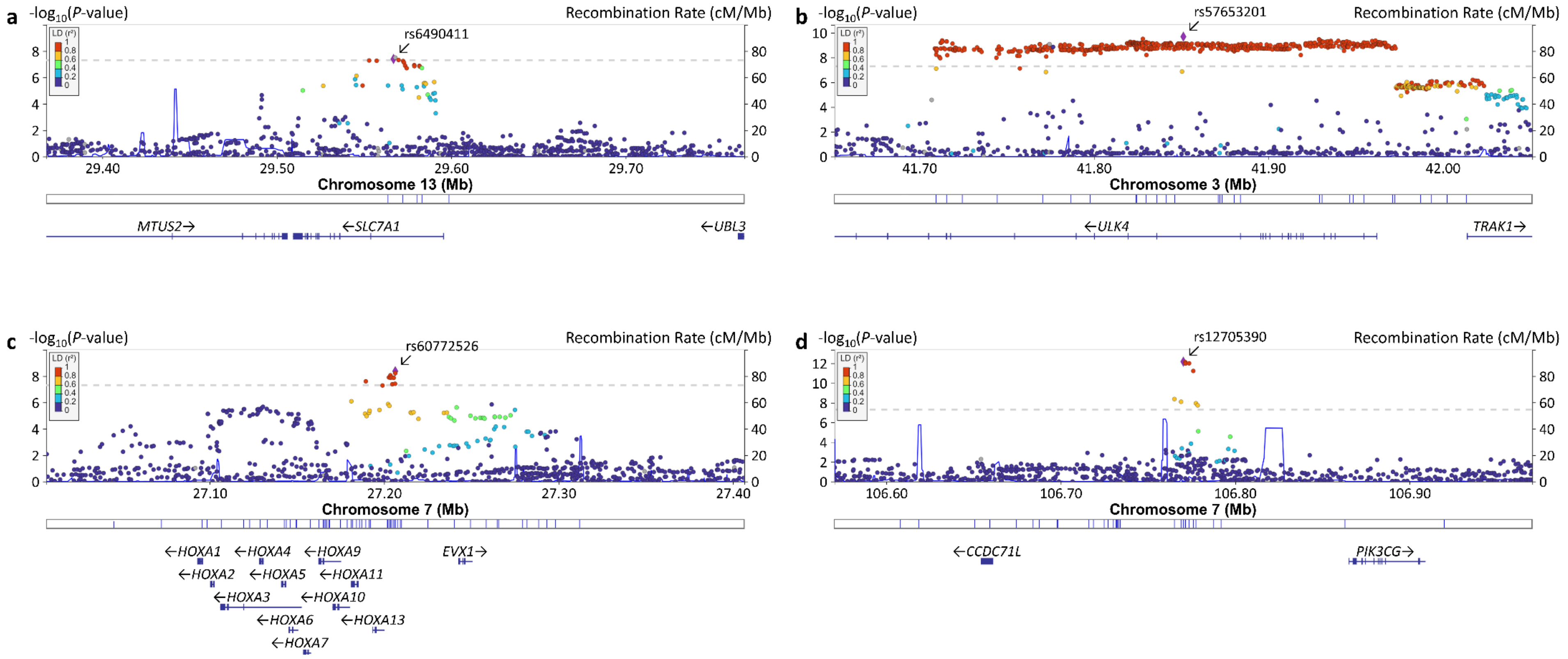

**Figure S4. Continued.** (e) rs2854746 and *IGFBP3* variants associated with PP in the total sample, (f) rs76622769 and *MCTP2* variants associated with SBP in the clinic-based cohorts, (g) rs2854746 and *IGFBP3* variants associated with PP in the clinic-based cohorts, and (h) rs12705390 and *PIK3CG* variants associated with PP in the prospective cohorts. A purple diamond indicates the top-ranked SNP at each locus. SNPs are color-coded according to their LD ($r^2$) with the top-ranked SNP in the region. Horizontal dotted lines represent the GWS threshold of $P$=5×10$^{-8}$. Vertical blue lines indicate locations of the high recombination rate among SNPs at the chromosomal position. Approximate location, transcription direction, and coding portions (exons represented by vertical bars) of genes are shown below the diagram. Mb=megabase, cM=centimorgan.

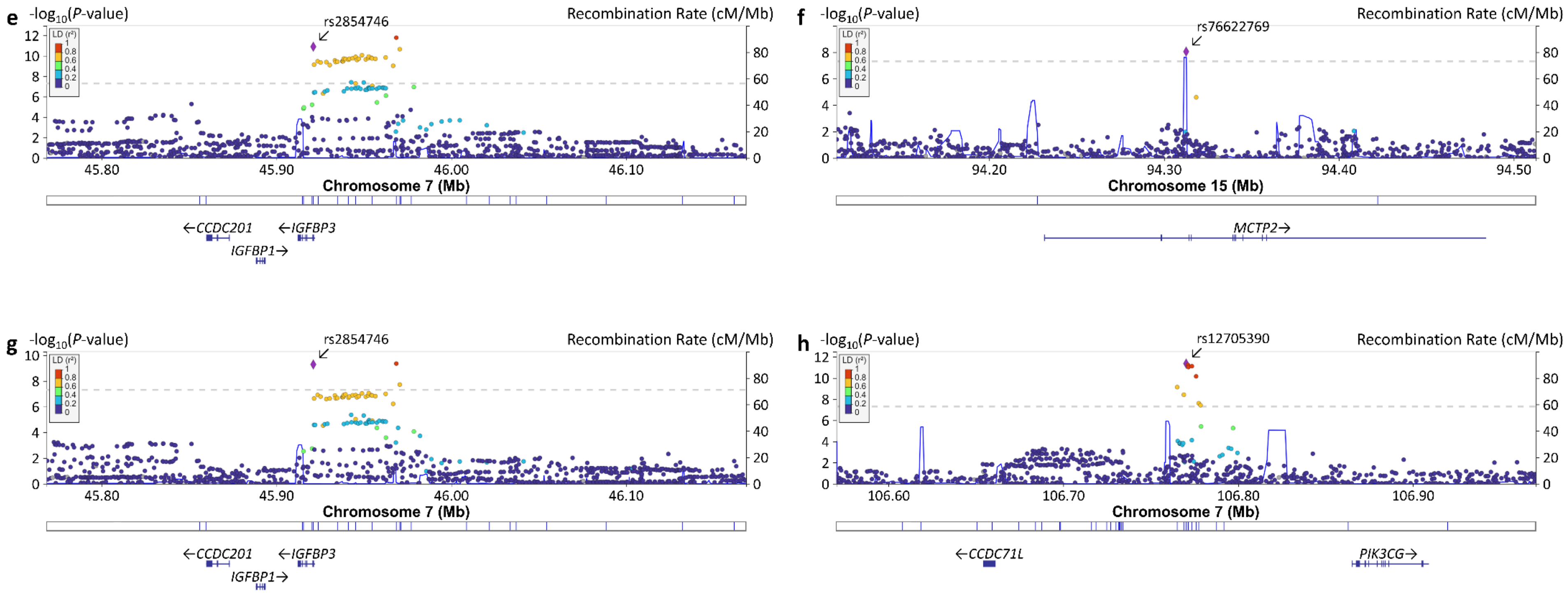

**Figure S5. Manhattan and QQ plots for genome-wide pleiotropy analyses**

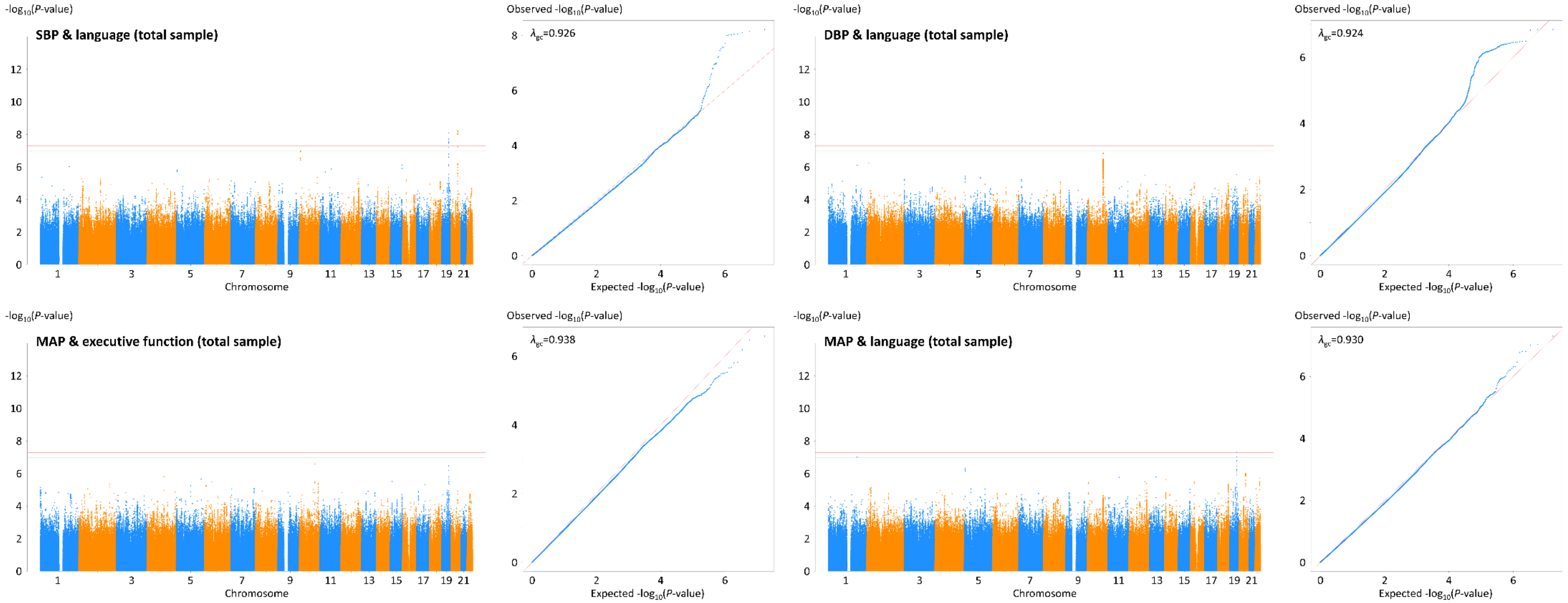

**Figure S5. Continued**

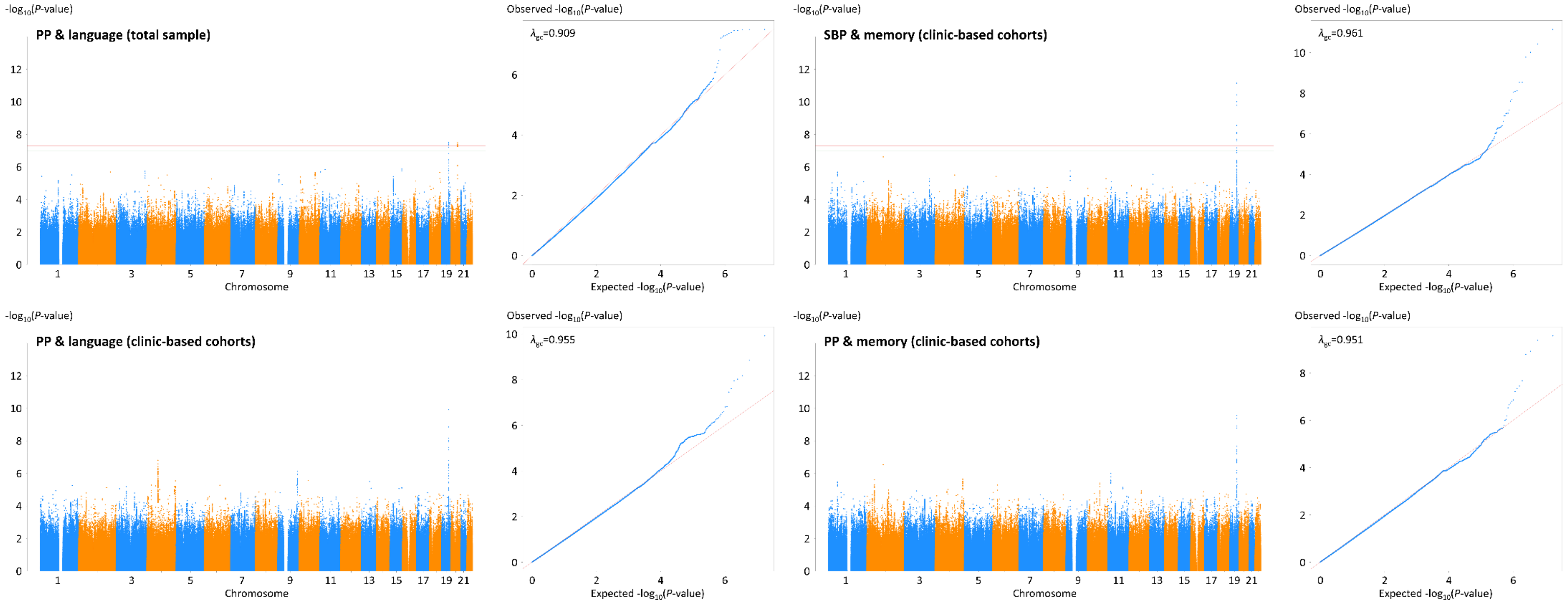

#### Figure S5. Continued

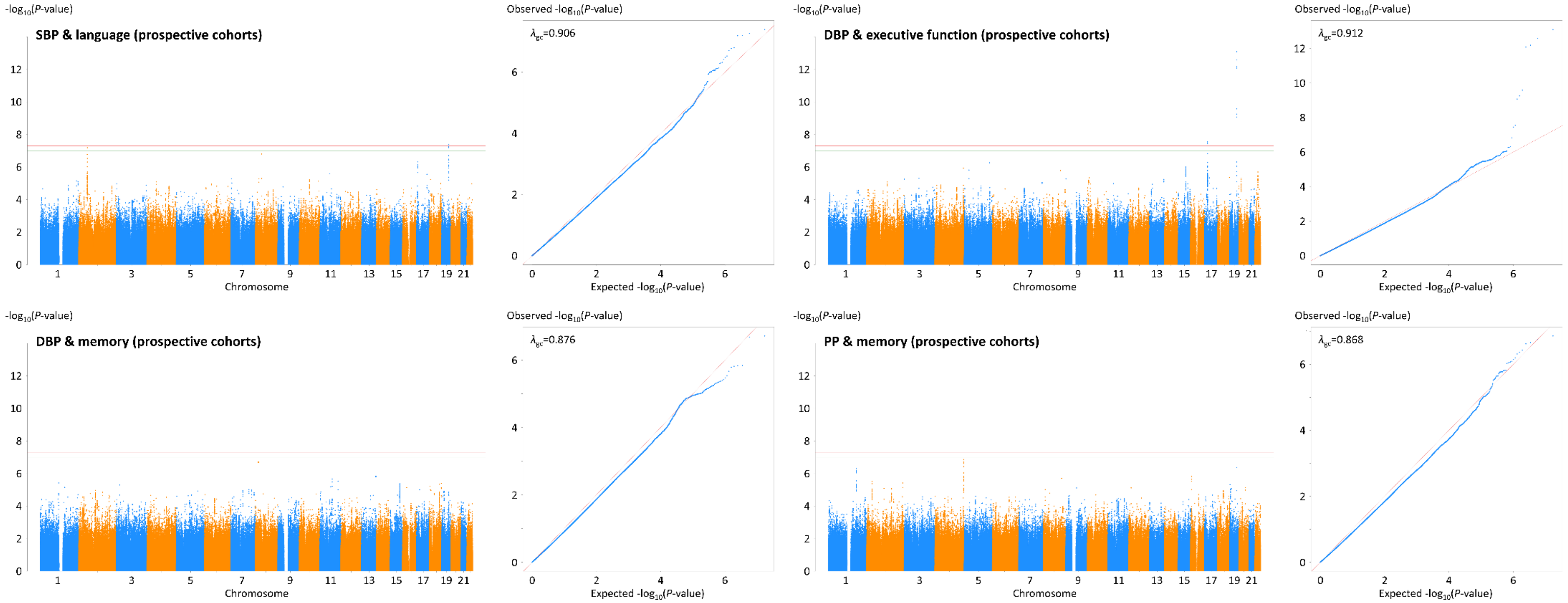

**Figure S6. LocusZoom plots for top-ranked pleiotropic variants identified in intergenic regions.** (a) rs73050834 and *LOC105371656* variants showing pleiotropy between MAP and language in the total sample, (b) rs117854720 and *PAX2* variants showing pleiotropy between MAP and executive function in the total sample, (c) rs10201413 and *LINC02946* variants showing pleiotropy between SBP and memory in the clinic-based cohorts, (d) rs10201413 and *LINC02946* variants showing pleiotropy between PP and memory in the clinic-based cohorts, and (e) rs57127265 and *LOC100128993* variants showing pleiotropy between DBP and memory in the prospective cohorts. A purple diamond indicates the top-ranked SNP at each locus. SNPs are color-coded according to their LD ($r^2$) with the top-ranked SNP in the region. Horizontal dotted lines represent the GWS threshold of $P$=5×10$^{-8}$. Vertical blue lines indicate locations of the high recombination rate among SNPs at the chromosomal position. Approximate location, transcription direction, and coding portions (exons represented by vertical bars) of genes are shown below the diagram. Mb=megabase, cM=centimorgan.

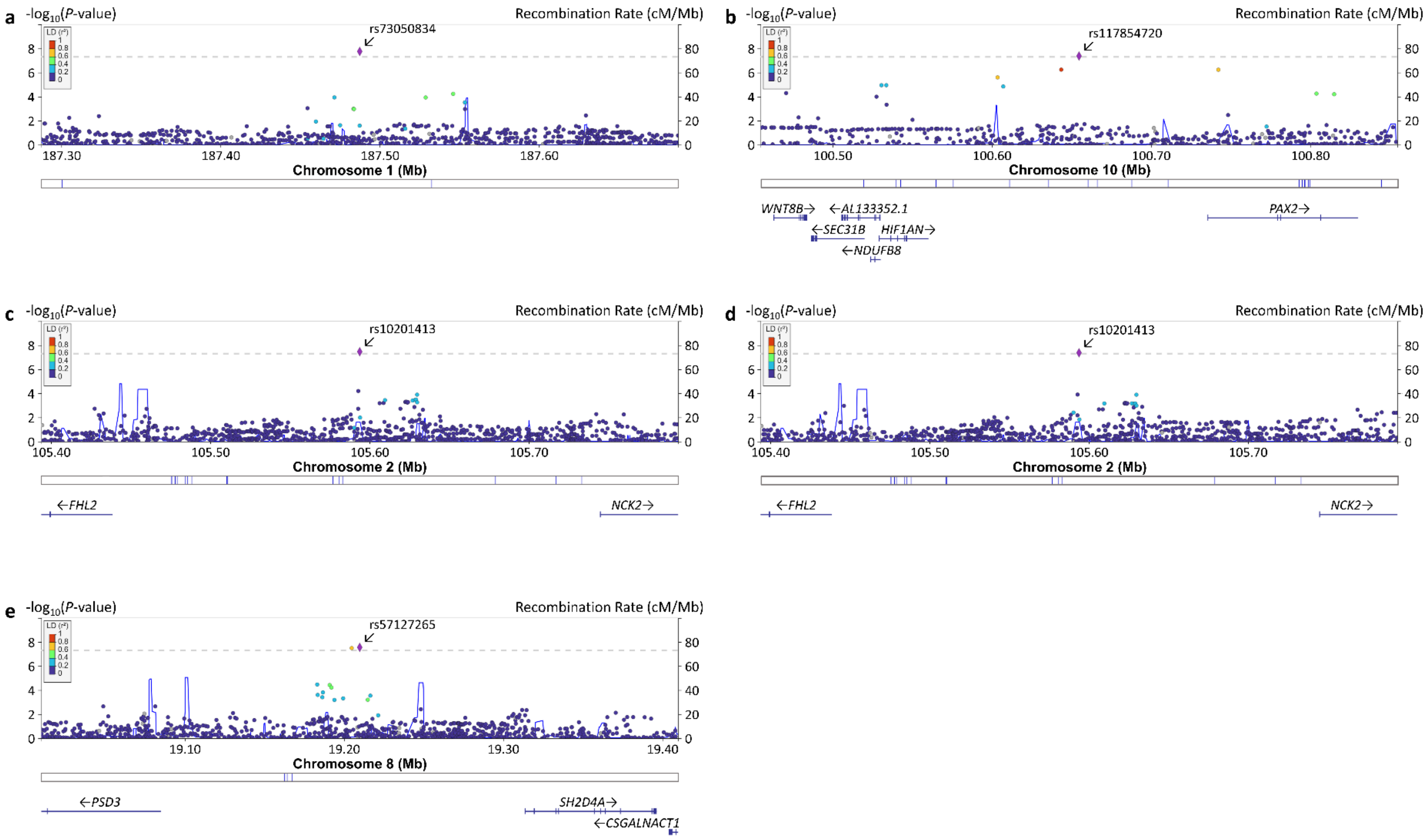
